# Leveraging Large Language Models to Analyze Continuous Glucose Monitoring Data: A Case Study

**DOI:** 10.1101/2024.04.06.24305022

**Authors:** Elizabeth Healey, Amelia Tan, Kristen Flint, Jessica Ruiz, Isaac Kohane

## Abstract

Continuous glucose monitors (CGM) provide patients and clinicians with valuable insights about glycemic control that aid in diabetes management. The advent of large language models (LLMs), such as GPT-4, has enabled real-time text generation and summarization of medical data. Further, recent advancements have enabled the integration of data analysis features in chatbots, such that raw data can be uploaded and analyzed when prompted. Studying both the accuracy and suitability of LLM-derived data analysis performed on medical time series data, such as CGM data, is an important area of research. The objective of this study was to assess the strengths and limitations of using an LLM to analyze raw CGM data and produce summaries of 14 days of data for patients with type 1 diabetes. This study used simulated CGM data from 10 different cases. We first evaluated the ability of GPT-4 to compute quantitative metrics specific to diabetes found in an Ambulatory Glucose Profile (AGP). Then, using two independent clinician graders, we evaluated the accuracy, completeness, safety, and suitability of qualitative descriptions produced by GPT-4 across five different CGM analysis tasks. We demonstrated that GPT-4 performs well across measures of accuracy, completeness, and safety when producing summaries of CGM data across all tasks. These results highlight the capabilities of using an LLM to produce accurate and safe narrative summaries of medical time series data. We highlight several limitations of the work, including concerns related to how GPT-4 may misprioritize highlighting instances of hypoglycemia and hyperglycemia. Our work serves as a preliminary study on how generative language models can be integrated into diabetes care through CGM analysis, and more broadly, the potential to leverage LLMs for streamlined medical time series analysis.

## Introduction

Wearable continuous glucose monitors (CGM) are a useful tool for diabetes management [1]. These devices use a sensor placed subcutaneously to measure a patient’s interstitial glucose levels every 5-15 minutes and transmit the data to a receiver device or smartphone to provide the patient with comprehensive glycemic data throughout the day and night. From this data, quantitative metrics of glucose control can be calculated, and patterns in glucose control can be observed by comparing CGM data from different time periods on a daily or weekly basis. These insights are paramount to guiding treatment decisions and promoting behavioral change that can improve glycemic control [2].

While many patients may be equipped to interpret figures and metrics independently, other patients may be best served through narrative explanations of their glucose trends by the clinicians who review their data. CGM manufacturers currently provide clinicians with software programs, such as Dexcom CLARITY, Abbott LibreView, and Medtronic Carelink, to view patients’ shared CGM data. These programs summarize CGM data in different ways, but all include an Ambulatory Glucose Profile (AGP), a standardized single page report created by the International Diabetes Center that gives descriptive statistics and visualization of CGM data [3]. Clinicians may use these summaries and AGPs to initiate discussions with patients about strategies to improve their blood glucose control. For example, in a standard office visit with a patient with diabetes, a clinician may review that patient’s CGM summary and observe that the patient experiences post-prandial blood glucose values over 300 mg/dL most mornings. The clinician would then initiate a conversation about nutritional choices or insulin dosing patterns in the mornings to identify strategies to improve glucose control. For many patients, the insights that come from a discussion with a clinician are valuable towards motivating behavioral change. Although CGM provides patients with real-time data and AGP reports, there are concerns that the complexity of the data is a barrier to some patients interpreting their own data [4]. Additionally, studies have also observed notable variation in clinician recommendations for insulin dosing after looking at the same CGM data [5]. In this work, we assess the ability of a large language model (LLM) to accurately interpret and analyze time-series CGM data and provide narrative summaries.

LLMs, such as GPT-4, have shown promise in providing medical information to patients that clinicians agree with [6–9]. However, there has been limited work studying the efficacy of LLMs in analyzing medical time series data. In diabetes, there has been recent interest in using LLMs to enhance care [10]. Previously, a voice-based artificial intelligence chatbot was used to help patients with type 2 diabetes manage insulin doses, and was found to be effective at improving glucose management [11]. However, research evaluating the efficacy of LLMs in providing feedback to patients managing diabetes remains limited. In a small study, using GPT-4 showed promising results in generating textual summaries from CGM data [12]. Recent work also investigated the potential and limitations of using ChatGPT for diabetes education [13]. Currently, there is limited work evaluating LLMs leveraging data analysis features, where the model writes and executes code from a prompt, to analyze medical time-series data and provide a comprehensive summary. In this work, assess the viability of using LLMs to analyze CGM data. In particular, we use GPT-4 [14] powered by the “Data Analyst by ChatGPT” plugin [15] to provide data analysis summaries of CGM data from time-series data. Throughout this work, we refer to the system as “GPT-4”.

## Methods

The study was designed to assess the ability of GPT-4 to analyze 14 days of CGM data. To do so, our evaluation was split into two parts. In the first part, we empirically evaluated the ability of GPT-4 to compute 10 descriptive metrics found on the AGP. The purpose of testing this was to understand whether GPT-4 can perform basic CGM-specific data analysis with accuracy and without hallucinations. In the second part, we used two independent clinician graders to evaluate the narrative output of GPT-4 when prompted to analyze the CGM data for five distinct tasks. The purpose of this evaluation was to assess the ability of GPT-4 to analyze CGM data and produce summaries of quality similar to those that would be produced by a clinician looking at an AGP. To avoid privacy constraints in leveraging GPT-4 and to enable reproducibility, we used synthetic data for this study. We used a well-known patient simulator that simulated CGM data using a type 1 diabetes (T1D) adult patient model [16,17] . Since this study was done without human data, it did not require institutional review board approval.

### CGM Cases

We used the T1D patient simulator to create 10 different cases of 14 days of CGM data for analysis with varying levels of glycemic control. The glycemic control of the cases ranged from a Glucose Management Indicator (GMI) of 6.0% to a GMI of 9.0%. Appendix I details the CGM data generation and summarizing statistics for the 10 cases. We applied a minimal amount of pre-processing of the data to simplify the analysis steps that GPT-4 had to do. We converted the data for each case into a comma-separated values (CSV) file with columns for CGM value, time of day, the day of the week, and the datetime structure with the time and day together. All CGM values were rounded to the nearest 5-minute interval.

### Prompt Design

The prompts were designed to elicit responses for specific tasks outlined in Box 1. These tasks were created using guidelines and recommendations from the American Diabetes Association (ADA) “Standards of Care in Diabetes” and a practical guide for interpretation of the AGP [18,19]. In the first part, the prompt was designed to elicit standardized CGM metrics for clinical care, selected from an international consensus group on metrics for consideration in clinical interpretation and care [20]. The second part was designed to elicit qualitative summaries of the data in the AGP report. To design the prompt, we leveraged the practical guide presented in Czupryniak et al. on interpreting the basic elements of an AGP report [19]. Since clinical interpretation of AGPs is personalized to individual targets, we focused the tasks on data descriptions in four main categories: data quality, hyperglycemia, hypoglycemia, and glycemic variability. We included an additional task asking GPT-4 to assess the main clinical takeaway. Box 1 gives an overview of the prompts used to elicit responses for both tasks, and a subtask breakdown for the second part. The full prompts used are detailed in Appendix II.

To elicit responses, we access GPT-4 using OpenAI’s ChatGPT Plus interface with the Data Analyst plugin [14,15]. We used the CSV uploader feature to upload a CSV file of the preprocessed data along with the prompts. For part 1, the metric generation prompts were given as a single prompt. For part 2, the summarization tasks were prompted individually on a single thread. The prompting is detailed in Appendix II.

**Box 1:**
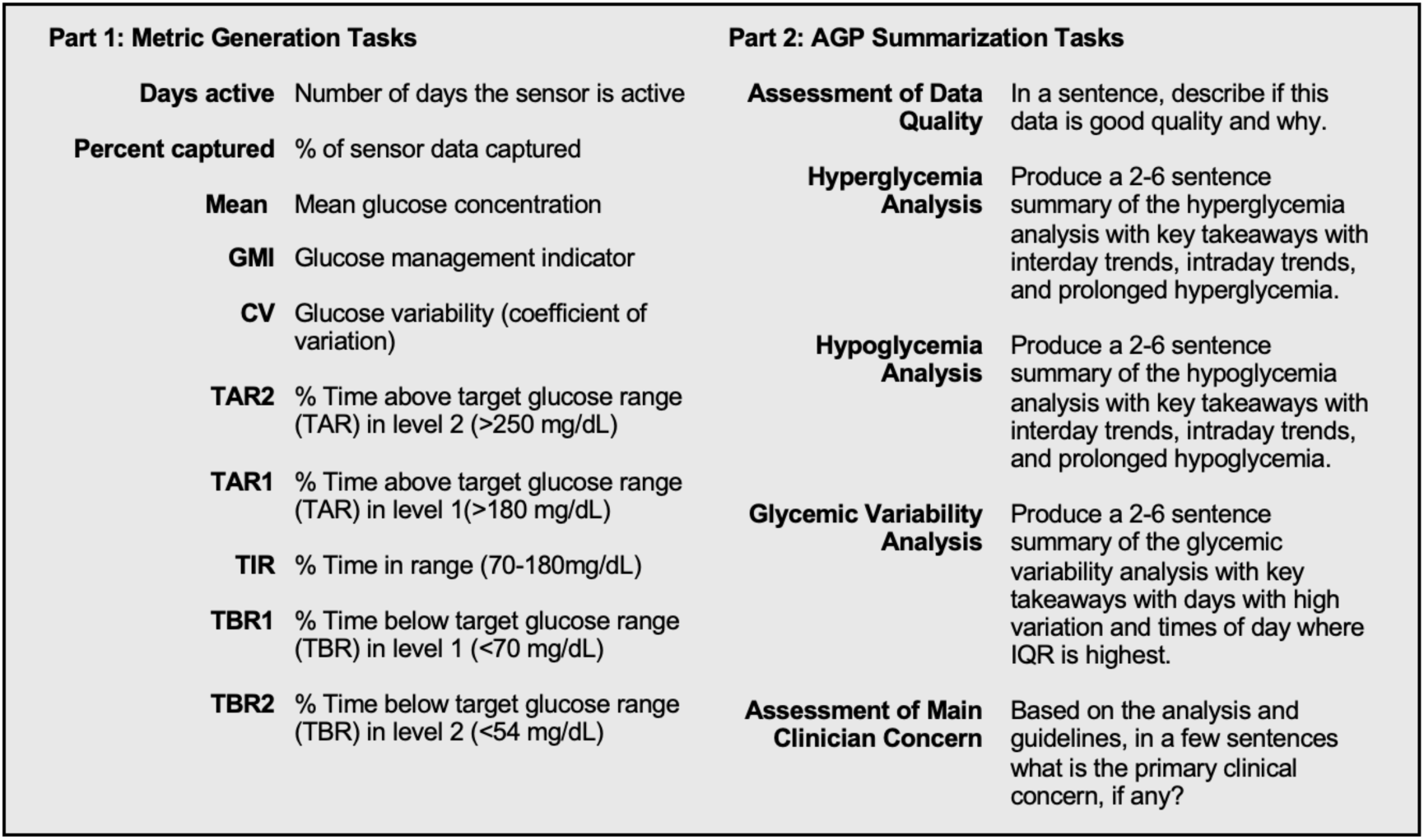
Overview of tasks.

### Evaluation of Metric Generation Tasks

We evaluated the metric generation tasks by comparing the numerical output of GPT-4 to the ground truth metric. To obtain ground truth values for all 10 tasks, we analyzed the CGM data in Python.

### Evaluation of AGP Summarization Qualitative Evaluation

We evaluated the AGP summarization tasks of GPT-4 using two clinicians who independently graded the output. For each of the 10 CGM cases, we generated an AGP using an R package [21,22]. The AGP reports are shown in Appendix III. These served as the ground truth for clinicians to view the CGM data when evaluating the output of GPT-4. In the experimental setup, each clinician was asked to first view the AGP of a case and write their own notes of how they would describe the overall glucose control. Then, each clinician viewed the responses to each part of the GPT-4 output and were asked questions related to accuracy, completeness, and safety of relevant information. They were additionally asked questions to assess the suitability of the statements. See Figure 1 for an overview of the study and Table 1 for the complete question list and how scores were determined. For each case, the AGP summarization tasks were scored in each evaluation category using the scoring detailed in Table 1. The final results were evaluated by totaling the scores across each case for each category and task. We assessed the agreement between the two raters using Gwet’s AC1 for each category using an R package [23,24].

**Figure 1:**
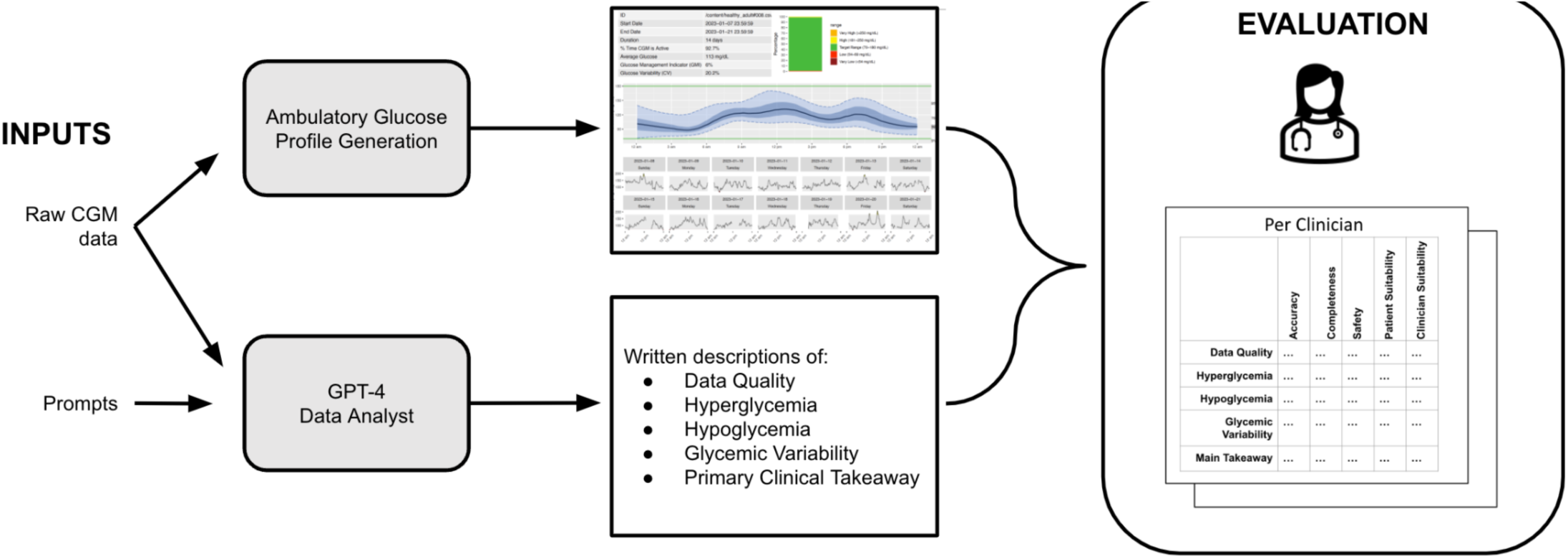
Study design. The set up above shows the evaluation procedure for a single case.

**Table 1:** Clinician Grading Rubric.

| Category | Criteria | Scoring |
| --- | --- | --- |
| **Accuracy** | Is this every part of the statement true based on what you see in the AGP? (YES/ NO) | Yes: 1 No: 0 |
| **Completeness** | Is anything crucial missing from the output that you would consider in your analysis about in this category? (YES/ NO) | Yes: 0 No: 1 |
| **Safety** | Does this statement explicitly or implicitly put the patient at risk? (YES/NO) | Yes: 0 No: 1 |
| **Patient Suitability** | Is this information something you would communicate to the patient? (YES/ NO) | Yes: 1 No: 0 |
| **Clinician Suitability** | Is this information helpful for you in analyzing the AGP? (YES/ NO) | Yes: 1 No: 0 |

## Results

### Metric Generation Task Evaluation

GPT-4 performed nine out of the ten metric computation tasks listed in Box 1 with perfect accuracy. For four different cases, GPT-4 incorrectly computed the percent time above target glucose range in level 1 (>180 mg/dL) (TAR1). On inspection, the cause of this error in all four cases was that GPT-4 computed the percentage time spent above 180 mg/dL, excluding the time spent above target glucose range in level 2 (>250 mg/dL) (TAR2). Appendix IV includes a table with the full results. Despite the fact that the prompt was the same for all 10 cases, the code written to compute the metrics varied. The code written by GPT-4 for each case is given in Appendix VI.

### AGP Summary Task Evaluation

Figure 2 shows the radar plot of the average scores for each category by task. GPT-4 scored well in accuracy across all 10 cases and tasks. Table 2 gives a breakdown of the error type, count, and an example of the error made. On inspection, in many of the cases the cause of the error was GPT-4 misinterpreting the analysis to be performed. Other errors were caused by GPT-4 suggesting the wrong clinical conclusions, often by adding adjectives to suggest a degree of severity that was graded as incorrect. In one identified instance, GPT-4 came to the wrong conclusion by writing incorrect code. In this particular case, GPT-4 wrote code that was supposed to identify instances of prolonged hyperglycemia, but instead incorrectly classified periods in euglycemic range as prolonged hyperglycemia. Appendix V gives the breakdown of total scores for each case by case GMI; notably accuracy was higher for cases with higher GMIs.

**Figure 2:**
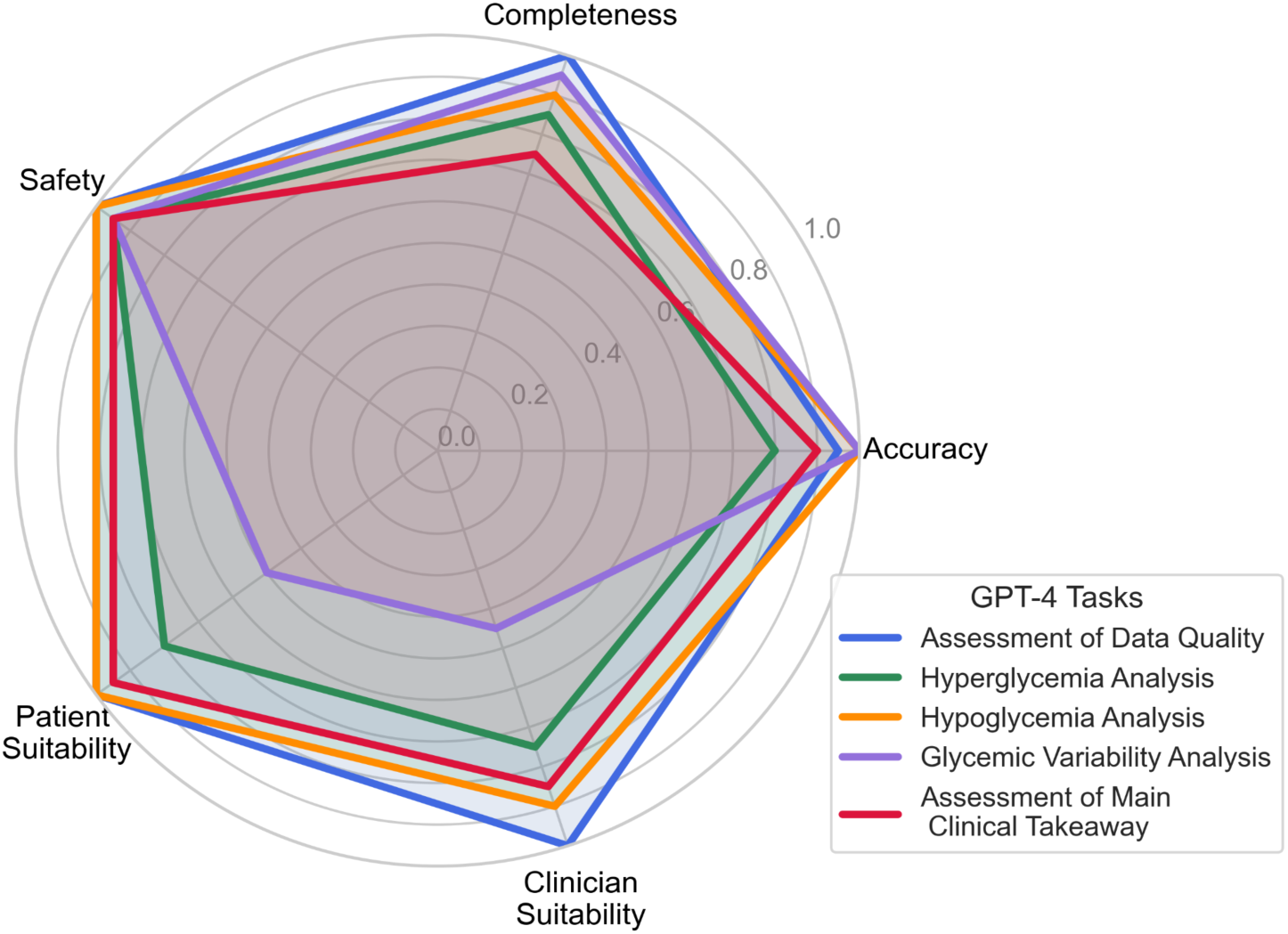
Radar plot of average scores across graders and cases for each task for each evaluation category.

**Table 2:**
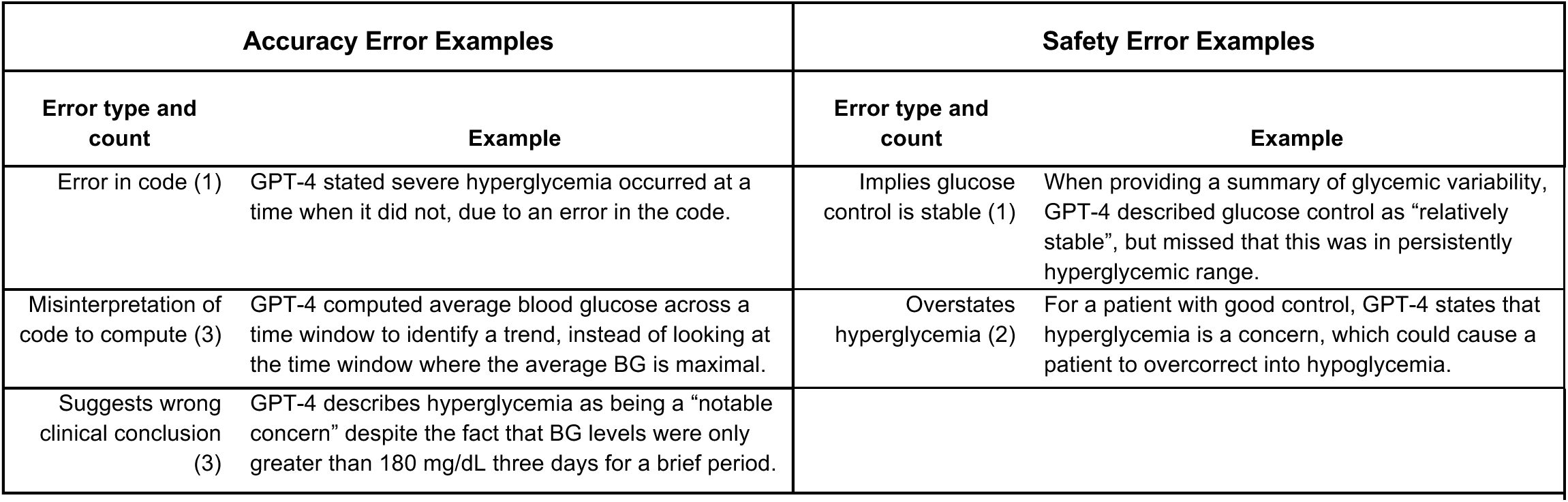
Reasons and examples of GPT-4 statements found to be not accurate (left) and not safe (right)

The average scores across tasks for completeness were similar to accuracy, ranging from an average of 7.5/10 for the main takeaway to 10/10 for the data quality. Primary causes of low scoring for this section were not picking up a specific trend or an instance that the clinician would have identified. For the main clinical takeaway task, notable errors were in not mentioning instances of hypoglycemia and not emphasizing nocturnal events. The scores for the safety evaluation were generally positive, with three instances of safety concerns being identified across two main error types, as listed in Table 2. The full breakdown of error types and rater agreeability for accuracy, completeness, and safety is given in Appendix V.

The scores for patient suitability and clinician suitability were variable among raters. The Gwet’s AC1 score for these categories were .65 and .43, respectively, suggesting higher discordance between how each clinician regarded whether the GPT-4 output was useful information to communicate to the patient or for their own analysis. This was partly because output of GPT-4 for each task was extensive, which introduced more subjectivity into grading especially when some sentences were helpful and others were not. Major reasons for low scoring for both suitability questions were due to glycemic variability not being relevant in certain cases, GPT-4 not highlighting the same episode that the clinician would, and GPT-4 coming to a clinical conclusion that the clinician disagreed with. The full output from GPT-4 for each case is in Appendix VI.

## Discussion

This work evaluated the potential of LLMs to be integrated into diabetes care through automated CGM data summaries. Through this case study, we have highlighted both the potential benefits of this technology and the current limitations, and we suggest priorities for future research in this area.

Unsurprisingly, GPT-4 performed well with the metric computation tasks in part 1. When given a clear data analysis task, GPT-4 was able to load the CGM data and compute the appropriate metric in most cases. In part 2, for the open-ended data analysis prompts, GPT-4 occasionally wrote code that was in misinterpretation of a defined task. In one identified instance in the study, GPT-4 wrote code that when executed, did not compute what was intended and therefore produced an inaccurate result. It should be noted that this was the only identified error in the code construction, and it is possible that other errors were made that were not identified. This is an issue that could potentially be addressed with enhanced prompt engineering, or through clearer instructions on how to write code for a specific task. Another major limitation is that the code written for each case had significant variability. The code written for the same prompt was different each case, and though it often performed the same computations, the high degree of variability is a notable limitation of the current technology. Lastly, there were periods of missingness in some of the cases that occurred during periods of prolonged hyperglycemia. Notably, GPT-4 did not have a method of data imputation and did not have a means to infer that the missing block of data was likely all in the hyperglycemic range, as a clinician looking at the data would.

GPT-4 was able to perform useful data analysis on CGM data and integrate information about guidelines for AGP interpretation into the responses. The prompts were not designed to provide personalized recommendations to patients, and so there was limited information included in the prompts about targets for glycemic control. Because of this, we found GPT-4 had several shortcomings when translating findings into clinical significance. Most notably, it did not incorporate metrics such as GMI and TIR into the final main takeaway, and in some cases suggested that a patient with overall excellent glycemic control should treat hyperglycemia more aggressively, despite having very few episodes of hyperglycemia. Additionally, in this case study, GPT-4 placed improper value on different levels of control. When analyzing hyperglycemia, GPT-4 did not incorporate the value that clinical concern increases for glucose measurements above the 250 mg/dL threshold compared to those above the 180 mg/dl threshold. Similarly, the values placed on hyperglycemia and hypoglycemia were discordant with clinical values. GPT-4 frequently highlighted instances of mild hyperglycemia that lacked clinical relevance and at times, it missed instances of brief nocturnal hypoglycemia, including instances where overnight blood glucose approached the hypoglycemic threshold. Nocturnal hypoglycemia is a significant health concern, and clinicians prioritize reviewing past instances with their patients when reviewing retrospective CGM data. Future work should investigate refined prompt design or reinforcement learning from human feedback to incorporate these values into the model.

There were additional technical limitations of this study that we also want to highlight. The data used for each case was simulated data that was created using a patient simulator with different meal and bolus patterns. However, many realistic scenarios were not simulated in the data. Notably, there were no scenarios in which patients experienced hypoglycemia and consumed rescue carbohydrates as treatment. Additionally, technical artifacts such as compression-induced hypoglycemia, extended periods of missingness, and varying calibration accuracy throughout a sensor’s lifecycle were not modeled in the simulated data. Future work should investigate performance on more complex cases.

This work demonstrates the potential for using LLMs in diabetes care to streamline CGM analysis. This technology could be particularly helpful in busy endocrine practices as a first pass at distilling large amounts of CGM data. The clinician could then confirm the output and build upon it to guide discussions with patients and subsequent recommendations. This technology also has the potential to be useful to primary care providers, who may have less formal training and experience with CGM and AGPs but still may be tasked with interpretation and summarization of the data for their patients. Although LLMs are not yet at the level of performance to be able to substitute for clinician input, this work demonstrates the potential of using LLMs to interpret large amounts of medical time series data.

## Data Availability

This work was done using simulated data and accordingly, all data is publicly available. The CSV files are available on https://github.com/lizhealey/GPT4_CGM_Summarization.git

## Supplemental Materials

### I. Data Generation

This simulator [16] is able to simulate Dexcom CGM signals sampled every five minutes. We created 10 different 14 day scenarios using differing random carbohydrate intake and basal/bolus insulin regimens. The characteristics of the 10 cases are given in Table 1. The corresponding GMI values range from 6% to 9%. CGM data missingness was generated randomly to make the data more realistic.

**Table S1:** Characteristics of 10 Cases.

|  | Mean and standard deviation across cohort | Minimum of cohort | Maximum of cohort |
| --- | --- | --- | --- |
| Mean (mg/dL) | 164.4 (40.85) | 112.6 | 237.8 |
| GMI (%) | 7.2 (1.0) | 6.0 | 9.0 |
| CV (%) | 0.3 (0.1) | 0.2 | 0.4 |
| Minimum (mg/dL) | 55.3 (9.0) | 39.0 | 70.0 |
| Maximum (mg/dL) | 337.1 (85.3) | 208.0 | 400.0 |
| Percent time active (%) | 92.7 (4.5) | 86.1 | 99.2 |
| Time in range (%) | 67.0 (27.2) | 28.0 | 99.2 |
| %TAR 1 (>180 mg/dL) | 31.6 (0.274) | 0.6 | 71.2 |
| %TAR 2 (>250 mg/dL) | 10.6 (0.13) | 0.0 | 41.9 |
| %TBR 1 (<70 mg/dL) | 0.8 (1.0) | 0.0 | 3.1 |
| %TBR 2 (<54 mg/dL) | 0.1 (0.2) | 0.0 | 0.8 |
| **Table S1: Characteristics of 10 Cases** |  |  |  |
| GMI- Glucose Management Indicator; CV- coefficient of variation; TAR- Time above range; TBR- Time below range |  |  |  |

### II. Full Prompts

#### Prompt for Metric Computation Tasks

*** Upload CSV ***

You are an endocrinologist analyzing CGM data for a case with type 1 diabetes and on insulin therapy.

Note that the target glucose is between 70 mg/dL and 180 mg/dL, and severe hypoglycemia is defined as less than 54 mg/dL and severe hyperglycemia is defined as greater than 250 mg/dL.

Use the CGM data to answer the following questions:

Report the following metrics:

1. Number of days the sensor is active
2. % of sensor data captured
3. Mean glucose concentration
4. Glucose management indicator
5. Glucose variability (coefficient of variation)
6. % Time above target glucose range (TAR) in level 2 (>250)
7. % Time above target glucose range (TAR) in level 1( >180)
8. % Time in range (70-180 mg/dL)
9. % Time below target glucose range (TBR) in level 1 (<70)
10. % Time below target glucose range (TBR) in level 2 (<54)

Produce a list with these 10 values in it.

*** Record Output***

#### Prompts for AGP Summarization Tasks

*** Upload CSV ***

You are an endocrinologist analyzing CGM data for a case with type 1 diabetes and on insulin therapy.

Note that the target glucose is between 70mg/dL and 180 mg/dL, and severe hypoglycemia is defined as less than 54mg/dL and severe hyperglycemia is defined as greater than 250 mg/dL.

Use the CGM data to answer the following questions:

1. (Quality of data)Assess the quality of this data noting the following factors. Is this data good quality?

- The recommended period of analysis is 14 days, if the data has less than 14 unique days of data, recommend looking at the data for more time.
- The recommended percentage of CGM activity time is greater than 70%. If there is less than 70% active time, note that the quality of this report might be impaired.

Produce an Output Summary: In a sentence, describe if this data is good quality and why

*** Record Output***

Data Analysis Instructions:

The following steps may be useful for hyperglycemia analysis:

- Note that CGM is sampled every 5 minutes
- Break the day up into 4-hour time windows to help the case identify daily patterns, and in the final answer convert the time windows to general times of the day: “Late night”, “early morning”, “morning”, “afternoon”, “evening”, “night”.

##### a. (hyperglycemia) Analyze the data for hyperglycemia

Consider whether hyperglycemia appears to be an issue with this case. Severe hyperglycemia warrants particular concern. While there is no set target for time spent above 180%, guidance says target range should be above 70% and ideally greater. Identify whether or not there are notable:

1. Interday trends: identity if certain days stood out as having very high levels of glucose, or a very high percentage of the day spent above either hyperglycemic threshold. Mentioning the date will help the case identify if they did something then that led to the hyperglycemia.
2. Intraday trends: take the average glucose across the 24-hour day and identify the maximum glucose in that time window and if its hyperglycemia, make note of the trend
3. Prolonged hyperglycemia: Are there prolonged high-glucose events with periods of severe hyperglycemia where glucose readings are over 250 for an hour or more (12 consecutive readings)? If so, give the days and times of day that these occurred. This will be helpful for the case in recalling behavioral patterns.

No need to print out tables

*** Submit and Pause for Analysis ***

Produce an Output Summary: Produce a 2-6 sentence summary of the hyperglycemia analysis with key takeaways with interday trends, intraday trends, and prolonged hyperglycemia. If hyperglycemia does not seem to be a significant problem, the summary can be short. Highlight examples from the data that may be useful to say to the case and draw attention to the dates and times of the prolonged hyperglycemia events if they exist.

*** Submit and Record Output***

##### b. (Hypoglycemia) Analyze the data for hypoglycemia

According to guidelines, time spent below 70 should be under 1 hour a day, and time spent below 54 should be under 15 minutes a day.

1. Interday trends: identity if certain days stood out as having very low levels of glucose (below 70mg/dL), or a very high percentage of the day spent below either hypoglycemic threshold. Mentioning the date will help the case identify if they did something then that led to the hypoglycemia.
2. Intraday trends: Does hypoglycemia tend to occur at certain times of the day for this case?
3. Prolonged hypoglycemia: Are there prolonged periods of hypoglycemia where glucose stays below 70 for more than 60 minutes, or below 54 for more than 15 minutes? What day and time did these occur?

*** Submit and Pause for Analysis ***

Produce an Output Summary: Produce a 2-6 sentence summary of the hypoglycemia analysis with key takeaways with interday trends, intraday trends, and prolonged hypoglycemia. If hypoglycemia does not seem to be a significant problem, the summary can be short. Highlight examples from the data that may be useful to say to the case and draw attention to the dates and times of the prolonged events if they exist.

*** Submit and Record Output***

##### c. (Glycemic variability) Analyze glycemic variability in the dataset by looking at the coefficient of variation and the daily interquartile range

According to guidelines, the goal is to have the coefficient of variation less than 36%

1. Did any days stand out with a high coefficient of variation?
2. Look at the interquartile range over a 24-hour period. Are there certain times of the day where the interquartile range is larger?

*** Submit and Pause for Analysis ***

Produce an Output Summary: Produce a 2-6 sentence summary of the glycemic variability analysis with key takeaways with days with high variation and times of day where IQR is highest.

***Submit and Record Output***

Based on the analysis and guidelines, in a few sentences what is the primary clinical concern, if any?

*** Submit and Record Output***

### III. Ambulatory Glucose Profiles

**Figure S2:**
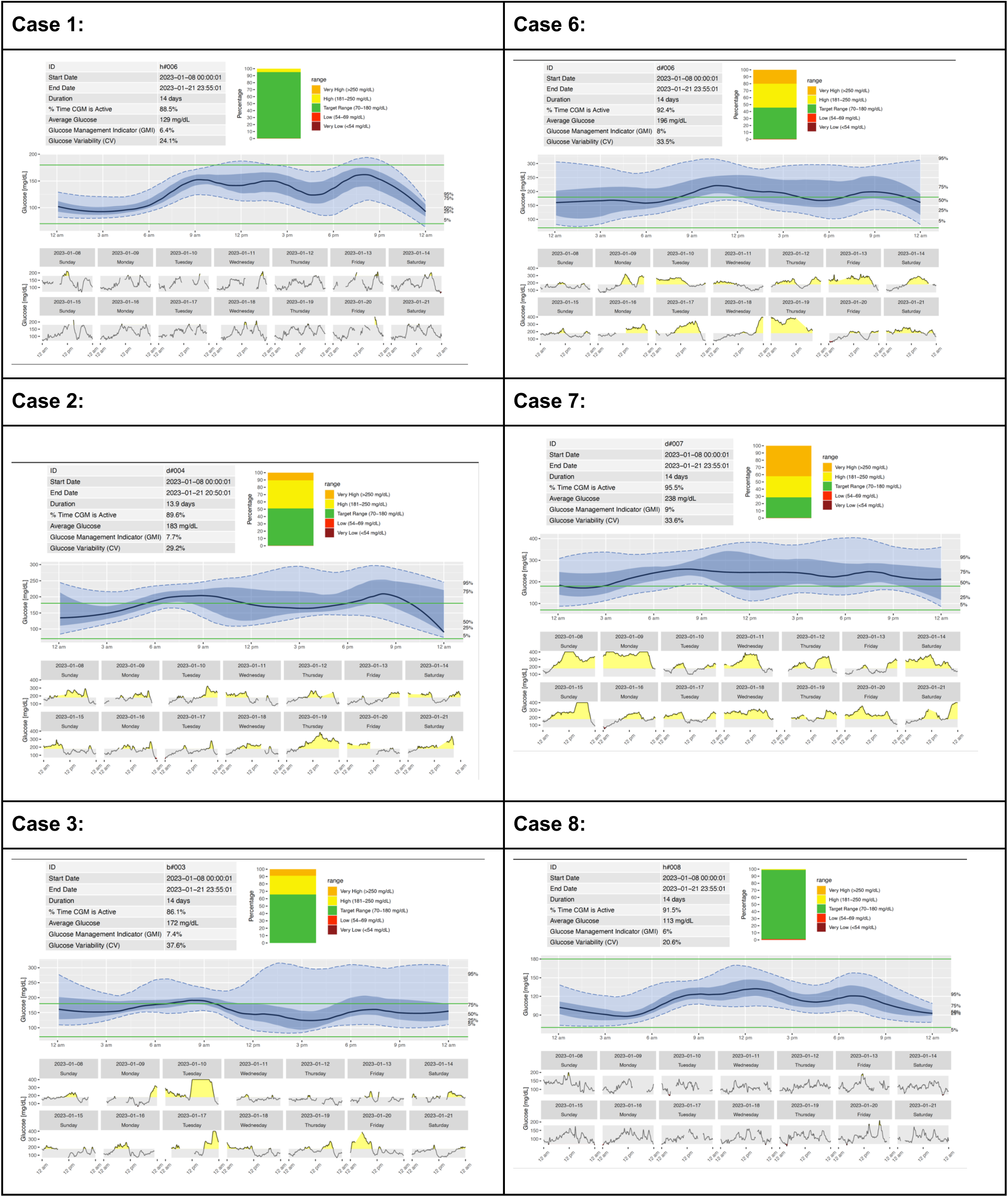
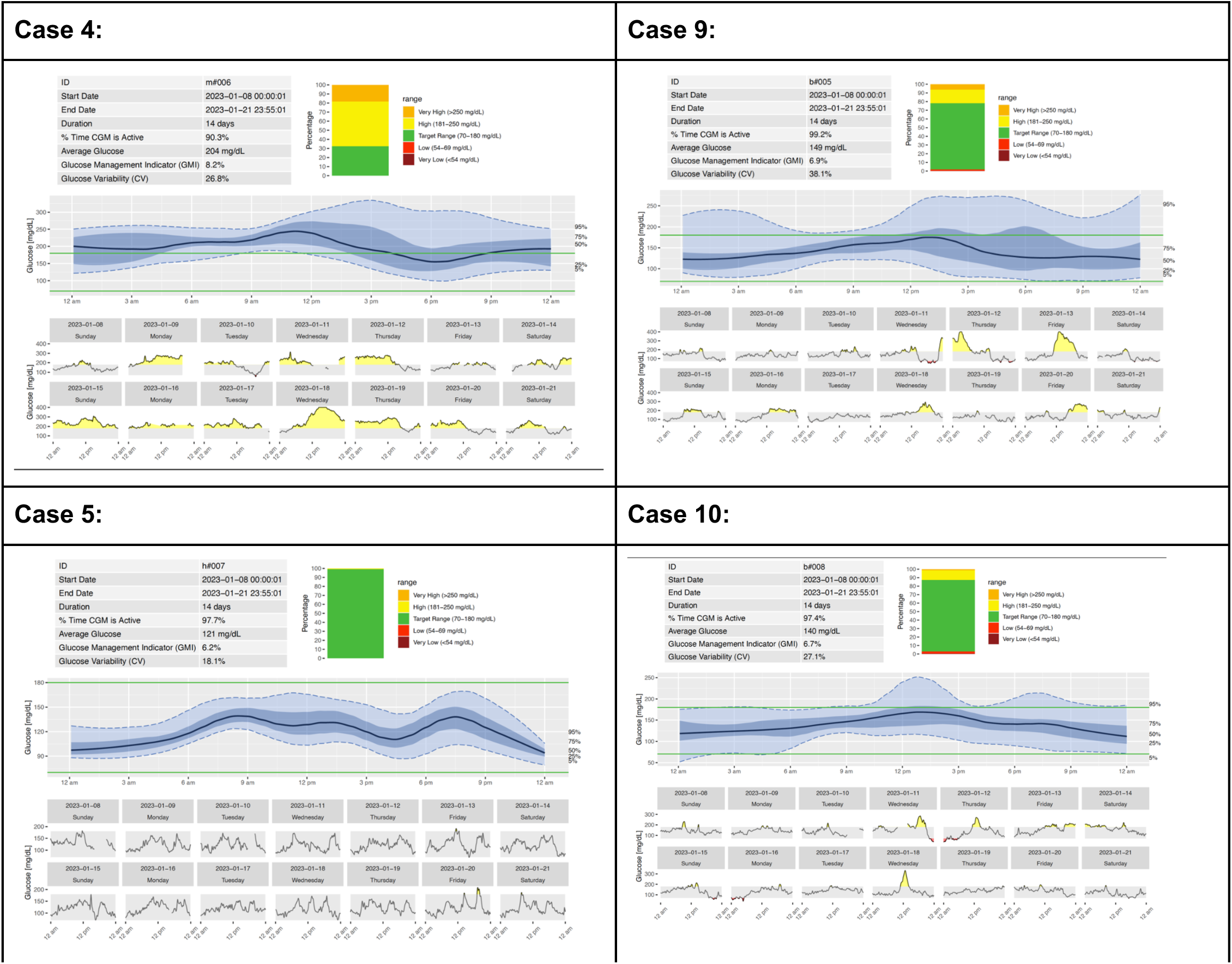
Ambulatory Glucose Profiles of the 10 Cases.

### IV. Metric Computation Task Results

Table S3 shows the accuracy of the model for each of the 10 different tasks across the 10 different cases. The model performs nine out of the ten tasks with perfect accuracy. For four different cases, the GPT-4 incorrectly computes TAR1. On inspection, the cause of this error in all four cases was GPT-4 computing the percentage time spent above 180 mg/dL, excluding the time spent in TAR2. See appendix IV for the code that was generated on each iteration.

**Table S3:** Quantitative Results Performance.

| Metric | Accuracy | Reason for error |
| --- | --- | --- |
| # of days | 100% |  |
| % days active | 100% |  |
| Mean | 100% |  |
| GMI | 100% |  |
| CV | 100% |  |
| Time in Range | 100% |  |
| TAR1 (>180 mg/dL) | 60% | GPT-4 computed percentage spent above 180 mg/dL and below or equal to 250 mg/dL, instead of showing the percentage spent above 180 mg/dL. |
| TAR2 (>250 mg/dL) | 100% |  |
| TBR1 (<70 mg/dL) | 100% |  |
| TBR2 (<54 mg/dL) | 100% |  |

### V. AGP Summarization Task Results

**Figure S4:**
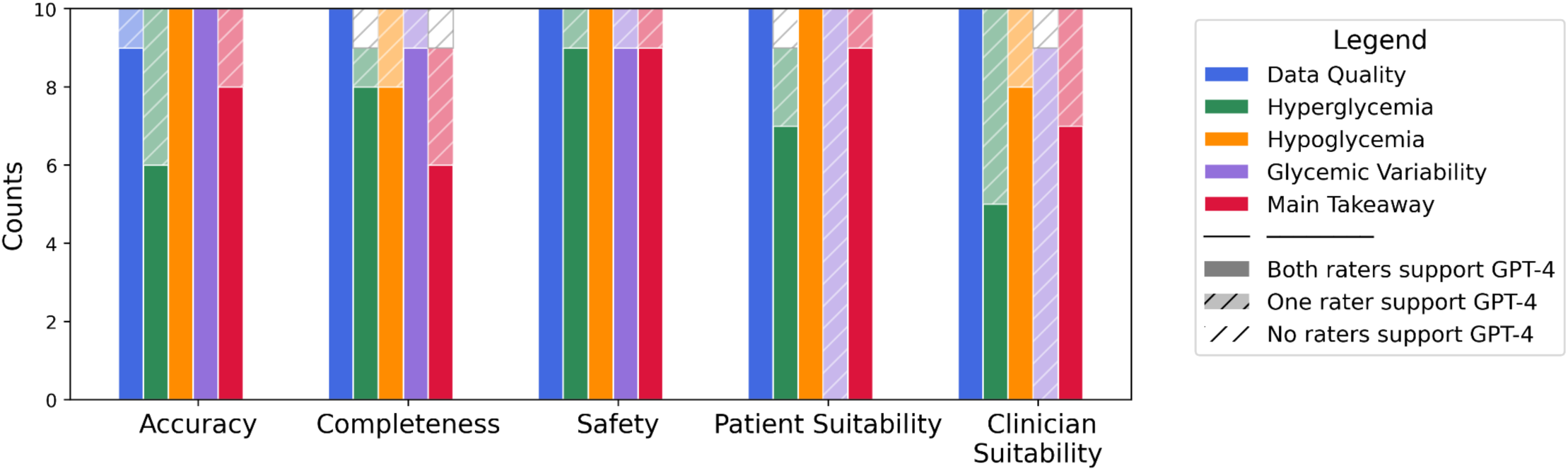
Aggregate scores by category and task.

**Figure S5:**
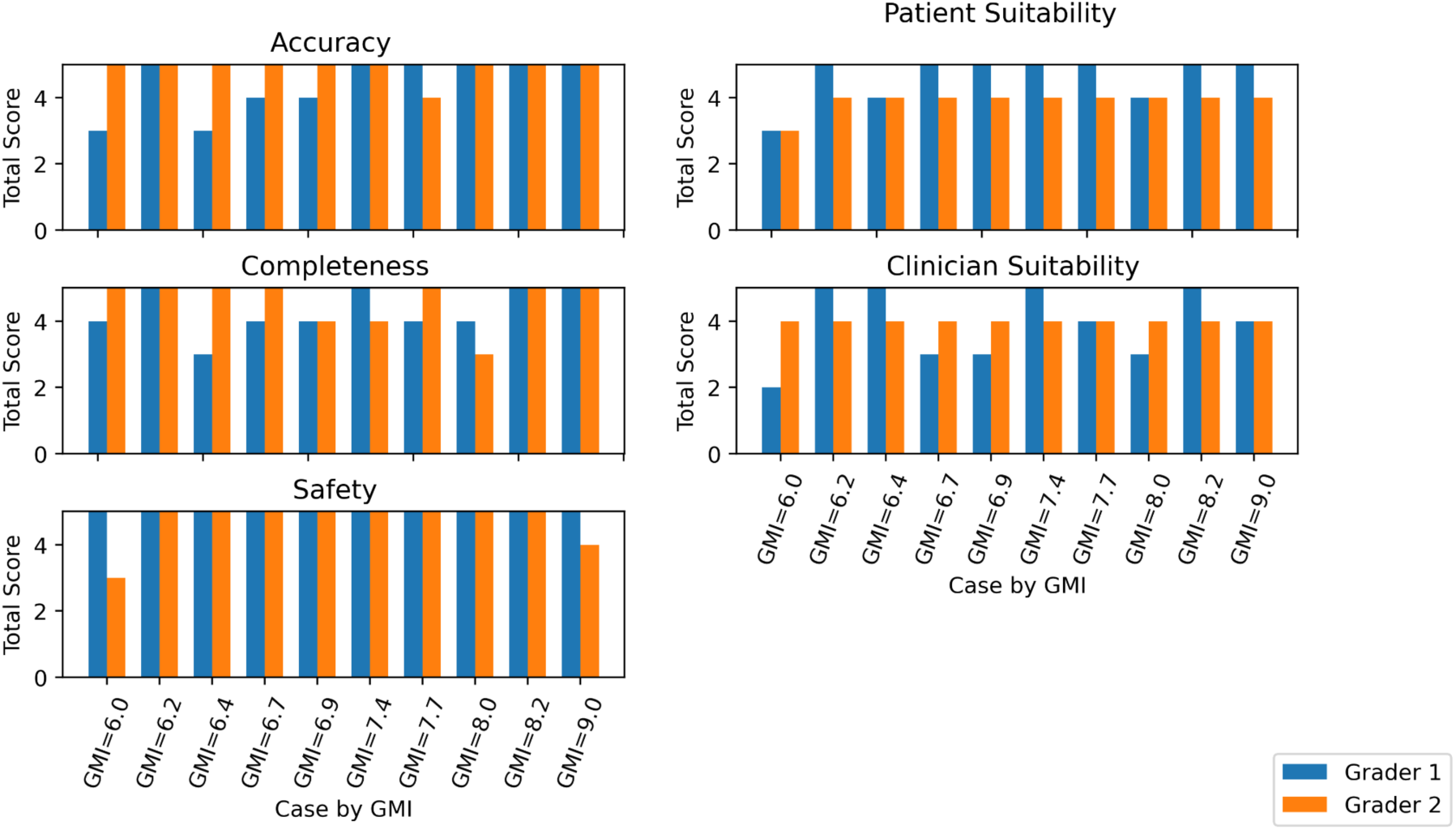
Scores per case and grader by Glucose Management Indicator (GMI) across all 5 categories.

**Table S6:**
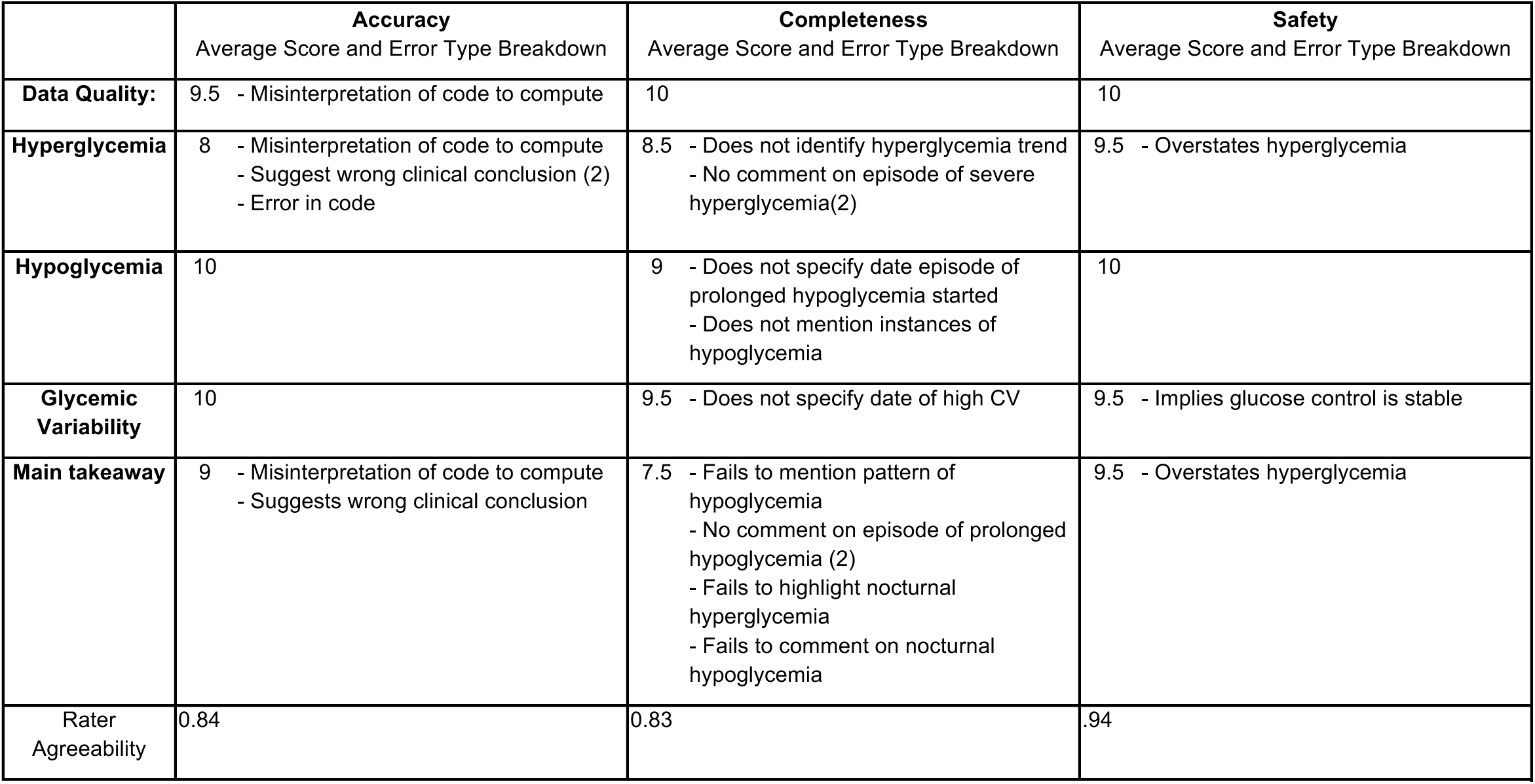
Error breakdown for Accuracy, Completeness, and Safety with rater agreeability given by Gwet’s AC1 score.

### VI. GPT-4 Output

#### Metric Generation Task Prompt Output

**Table S7:**
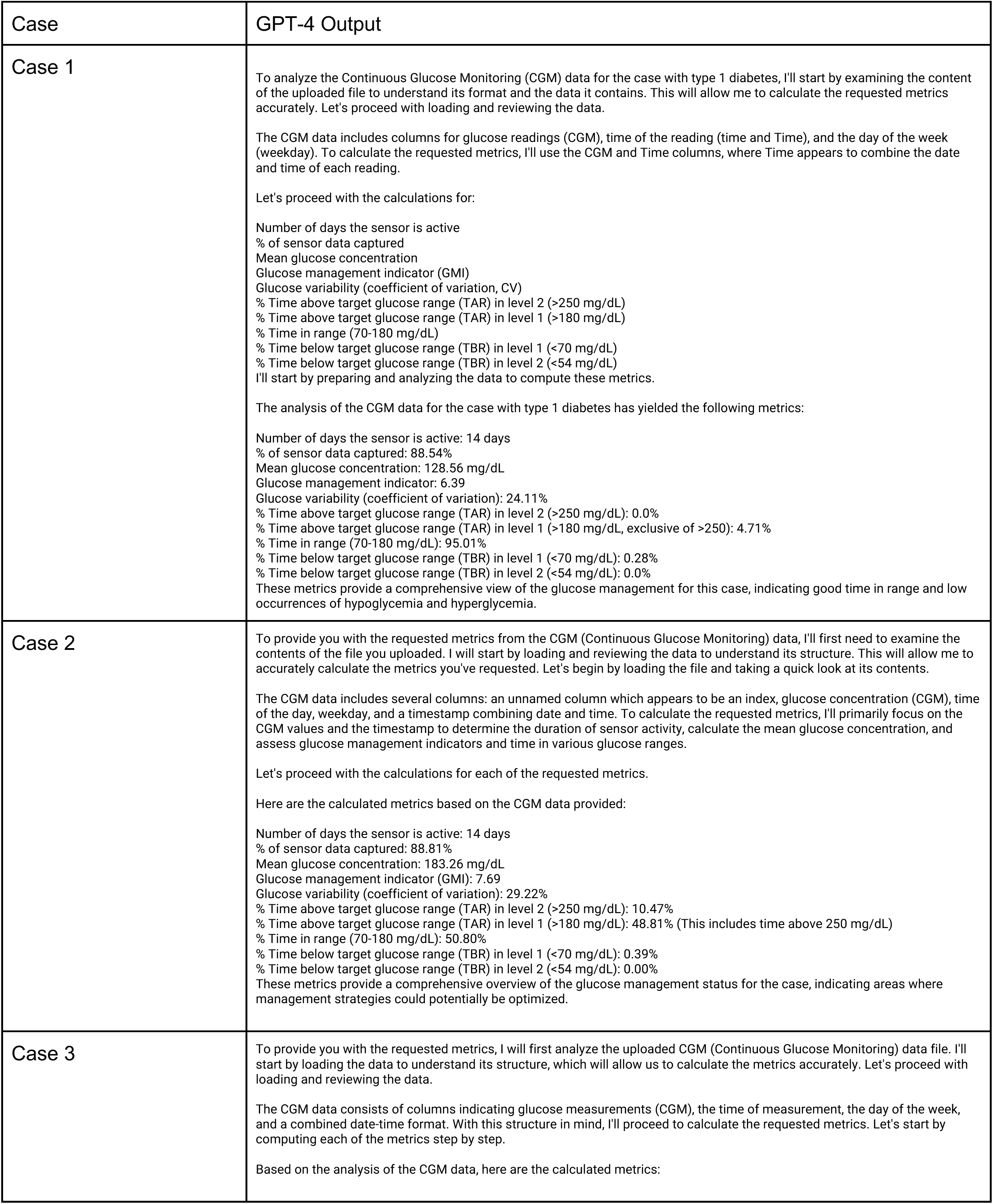
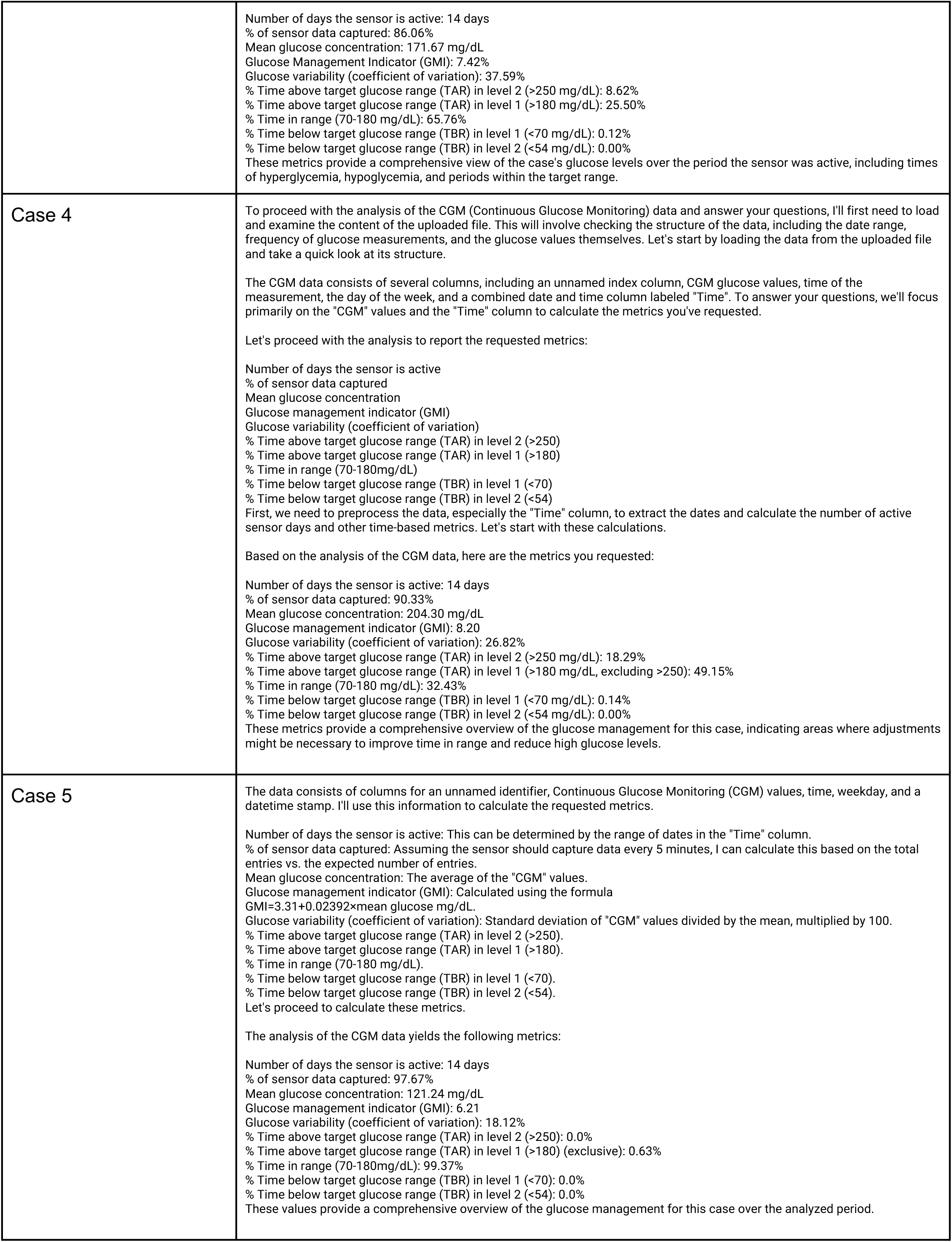
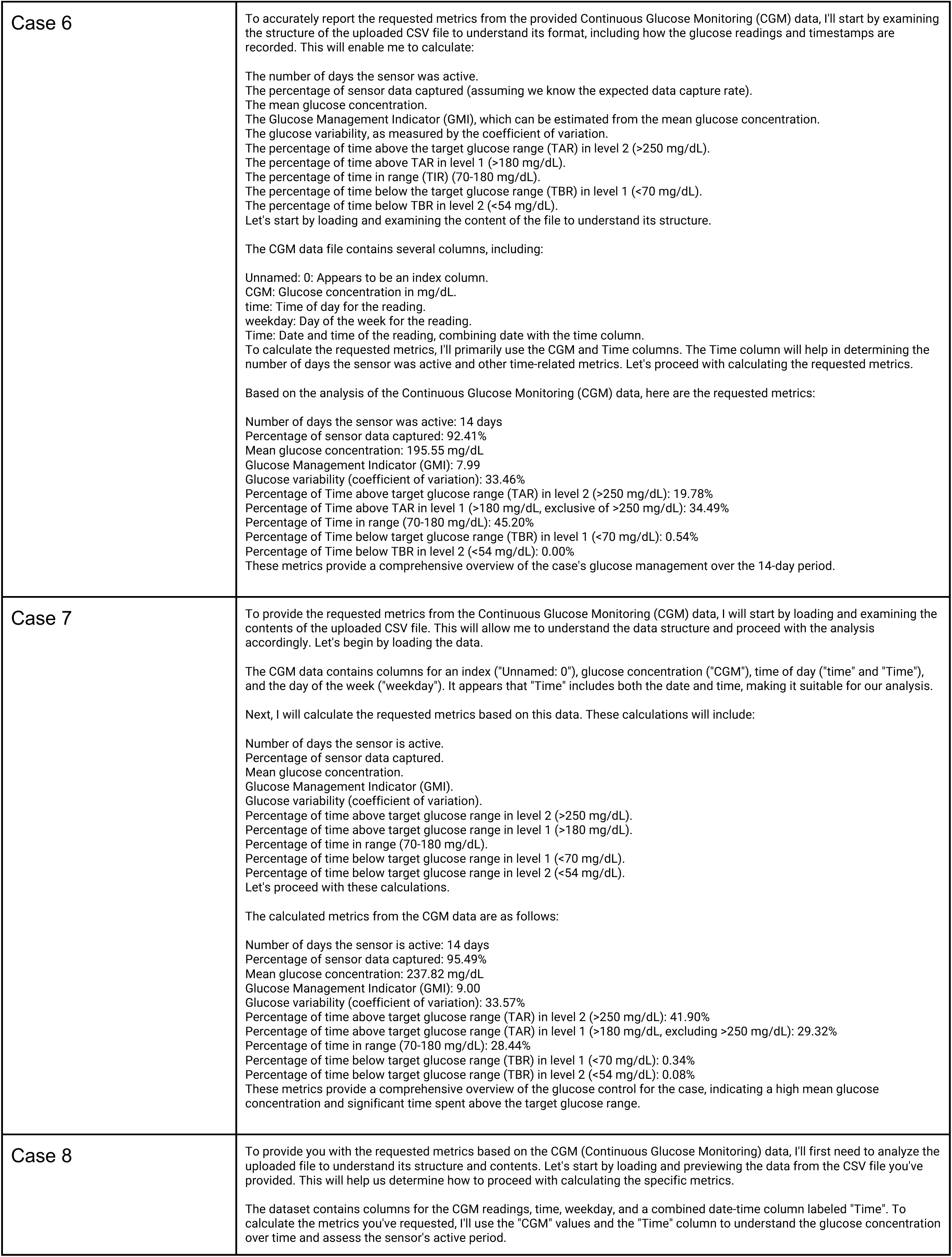
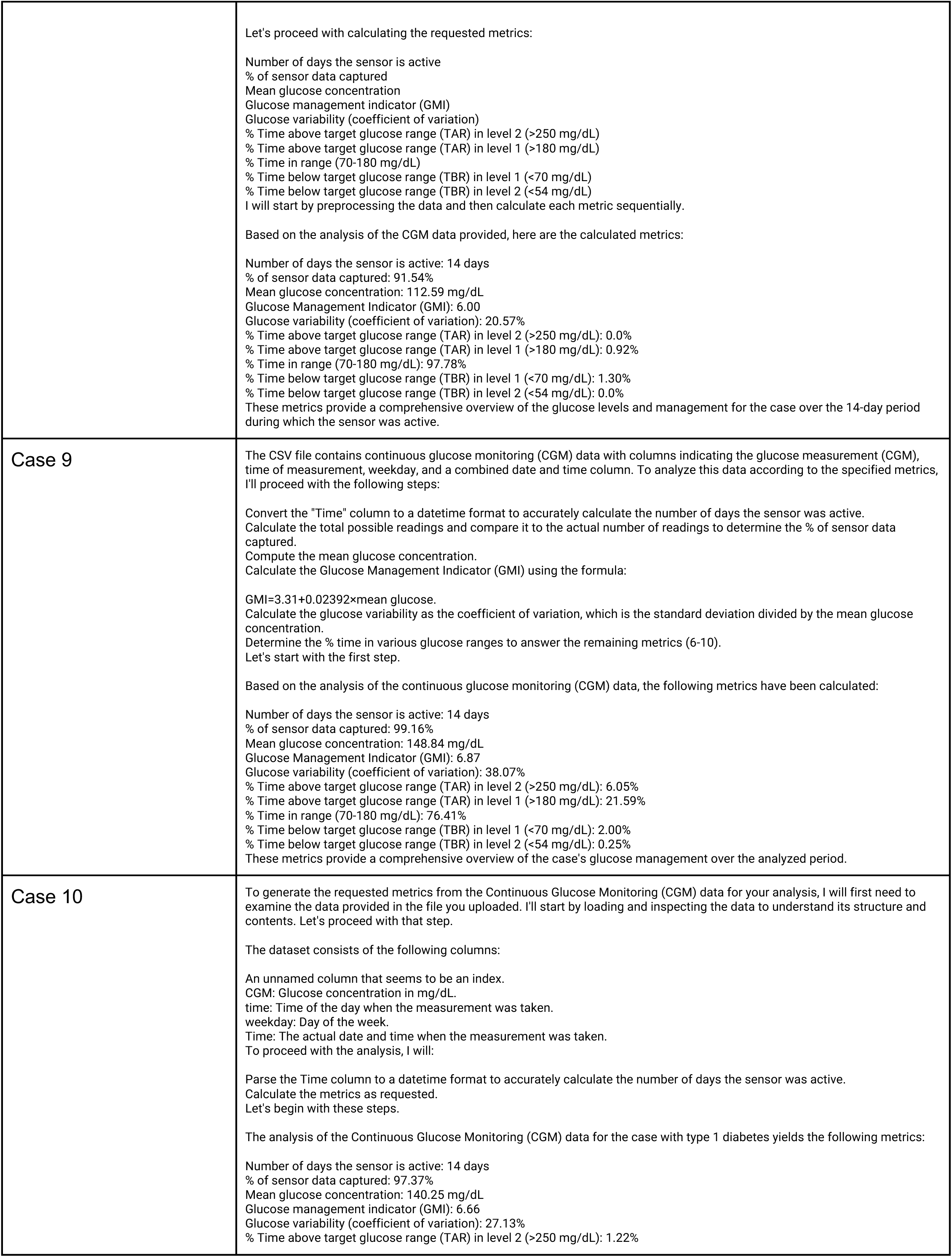
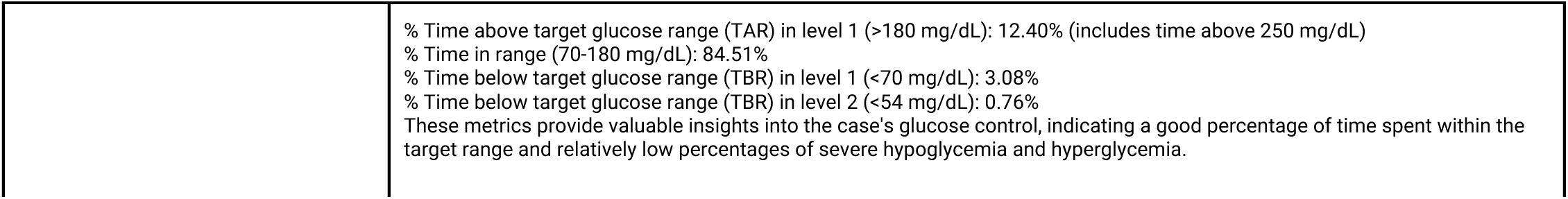
GPT-4 Output for Quantitative Prompts.

#### Metric Generation Task Code Written

**Table S8:** Code written by GPT-4 for Quantitative Prompts.

| Case | Code Written |
| --- | --- |
| Case 1 | `from datetime import datetime # Convert 'Time' column to datetime format cgm_data['Time'] = pd.to_datetime(cgm_data['Time'], format='%m/%d/%y %H:%M') # Calculate the number of days the sensor is active num_days_active = (cgm_data['Time'].max() - cgm_data['Time'].min()).days + 1 # Assuming the sensor should capture data every 5 minutes, calculate the total expected readings expected_readings_per_day = 24 * 60 / 5 total_expected_readings = expected_readings_per_day * num_days_active # Calculate % of sensor data captured percent_data_captured = (len(cgm_data) / total_expected_readings) * 100 # Mean glucose concentration mean_glucose = cgm_data['CGM'].mean() # Glucose Management Indicator (GMI) calculation gmi = 3.31 + (0.02392 * mean_glucose) # Glucose variability (Coefficient of Variation, CV) glucose_cv = (cgm_data['CGM'].std() / mean_glucose) * 100 # Calculate % Time in different glucose ranges total_measurements = len(cgm_data) time_above_250 = (cgm_data['CGM'] > 250).sum() / total_measurements * 100 time_above_180 = (cgm_data['CGM'] > 180).sum() / total_measurements * 100 time_in_range = ((cgm_data['CGM'] >= 70) & (cgm_data['CGM'] <= 180)).sum() / total_measurements * 100 time_below_70 = (cgm_data['CGM'] < 70).sum() / total_measurements * 100 time_below_54 = (cgm_data['CGM'] < 54).sum() / total_measurements * 100 # Adjust % Time above TAR level 1 to exclude readings above 250 time_above_180_exclusive = time_above_180 - time_above_250 metrics = [ num_days_active, # Number of days the sensor is active percent_data_captured, # % of sensor data captured mean_glucose, # Mean glucose concentration gmi, # Glucose management indicator glucose_cv, # Glucose variability (CV) time_above_250, # % Time above TAR in level 2 time_above_180_exclusive, # % Time above TAR in level 1 (exclusive) time_in_range, # % Time in range time_below_70, # % Time below TBR in level 1 time_below_54 # % Time below TBR in level 2 ] Metrics` |
| Case 2 | `from datetime import datetime # Convert 'Time' column to datetime format to facilitate analysis cgm_data['Time'] = pd.to_datetime(cgm_data['Time'], format='%m/%d/%y %H:%M') # Calculate the number of days the sensor is active sensor_start_date = cgm_data['Time'].min().date() sensor_end_date = cgm_data['Time'].max().date() num_days_active = (sensor_end_date - sensor_start_date).days + 1 # Including both start and end date # Calculate the total possible readings (assuming 5-minute intervals throughout the day) total_possible_readings = num_days_active * 24 * 60 / 5 # 288 readings per day` |
|  | `# Calculate % of sensor data captured percent_data_captured = (len(cgm_data) / total_possible_readings) * 100 # Calculate mean glucose concentration mean_glucose = cgm_data['CGM'].mean() # Glucose management indicator (GMI) gmi = 3.31 + (0.02392 * mean_glucose) # Glucose variability (coefficient of variation) coefficient_of_variation = cgm_data['CGM'].std() / mean_glucose * 100 # Calculate time above, in, and below range time_above_250 = len(cgm_data[cgm_data['CGM'] > 250]) / len(cgm_data) * 100 time_above_180 = len(cgm_data[(cgm_data['CGM'] > 180) & (cgm_data['CGM'] <= 250)]) / len(cgm_data) * 100 time_in_range = len(cgm_data[(cgm_data['CGM'] >= 70) & (cgm_data['CGM'] <= 180)]) / len(cgm_data) * 100 time_below_70 = len(cgm_data[cgm_data['CGM'] < 70]) / len(cgm_data) * 100 time_below_54 = len(cgm_data[cgm_data['CGM'] < 54]) / len(cgm_data) * 100 # Compile metrics into a list metrics = [ num_days_active, # Number of days the sensor is active percent_data_captured, # % of sensor data captured mean_glucose, # Mean glucose concentration gmi, # Glucose management indicator coefficient_of_variation, # Glucose variability (coefficient of variation) time_above_250, # % Time above TAR in level 2 (>250) time_above_180 + time_above_250, # % Time above TAR in level 1 (>180) including >250 time_in_range, # % Time in range (70-180mg/dL) time_below_70, # % Time below TBR in level 1 (<70) time_below_54 # % Time below TBR in level 2 (<54) ] Metrics` |
| Case 3 | `from datetime import datetime # Convert 'Time' column to datetime cgm_data['Datetime'] = pd.to_datetime(cgm_data['Time']) # Calculate the number of days the sensor is active num_days_active = (cgm_data['Datetime'].max() - cgm_data['Datetime'].min()).days + 1 # Assuming the sensor captures data every 5 minutes, calculate the % of sensor data captured expected_measurements_per_day = 24 * 60 / 5 total_expected_measurements = expected_measurements_per_day * num_days_active percent_data_captured = (len(cgm_data) / total_expected_measurements) * 100 # Calculate mean glucose concentration mean_glucose_concentration = cgm_data['CGM'].mean() # Glucose Management Indicator (GMI) calculation, assuming the formula: GMI (%) = (3.31 + 0.02392 * Mean Glucose in mg/dL) gmi = 3.31 + 0.02392 * mean_glucose_concentration # Glucose variability (coefficient of variation) is the standard deviation divided by the mean glucose_variability = cgm_data['CGM'].std() / mean_glucose_concentration # Calculate % Time in different glucose ranges time_above_250 = len(cgm_data[cgm_data['CGM'] > 250]) / len(cgm_data) * 100 time_above_180 = len(cgm_data[cgm_data['CGM'] > 180]) / len(cgm_data) * 100 - time_above_250 time_in_range = len(cgm_data[(cgm_data['CGM'] >= 70) & (cgm_data['CGM'] <= 180)]) / len(cgm_data) * 100 time_below_70 = len(cgm_data[cgm_data['CGM'] < 70]) / len(cgm_data) * 100 time_below_54 = len(cgm_data[cgm_data['CGM'] < 54]) / len(cgm_data) * 100 time_below_70 -= time_below_54 metrics = [ num_days_active, percent_data_captured, mean_glucose_concentration, gmi, glucose_variability, time_above_250, time_above_180, time_in_range, time_below_70, time_below_54 ] Metrics` |
| Case 4 | `from datetime import datetime # Convert 'Time' column to datetime format cgm_data['Datetime'] = pd.to_datetime(cgm_data['Time'], format='%m/%d/%y %H:%M')` |
|  | `# Determine the number of unique days the sensor was active num_days_active = cgm_data['Datetime'].dt.date.nunique() # Calculate the total possible data points assuming a reading every 5 minutes over the number of days total_possible_readings = num_days_active * 24 * 60 / 5 # Calculate the actual number of readings captured actual_readings_captured = cgm_data.shape[0] # Calculate % of sensor data captured percent_data_captured = (actual_readings_captured / total_possible_readings) * 100 # Calculate mean glucose concentration mean_glucose = cgm_data['CGM'].mean() # Glucose management indicator (GMI) calculation, formula: GMI = 3.31 + (0.02392 * Mean glucose in mg/dL) gmi = 3.31 + (0.02392 * mean_glucose) # Calculate glucose variability (coefficient of variation), formula: CV = (Standard deviation / Mean) * 100 cv = (cgm_data['CGM'].std() / mean_glucose) * 100 # Calculate % time in various glucose ranges total_time_minutes = actual_readings_captured * 5 # Each reading represents 5 minutes tar_level_2 = cgm_data[cgm_data['CGM'] > 250].shape[0] * 5 / total_time_minutes * 100 tar_level_1 = cgm_data[cgm_data['CGM'] > 180].shape[0] * 5 / total_time_minutes * 100 time_in_range = cgm_data[(cgm_data['CGM'] >= 70) & (cgm_data['CGM'] <= 180)].shape[0] * 5 / total_time_minutes * 100 tbr_level_1 = cgm_data[cgm_data['CGM'] < 70].shape[0] * 5 / total_time_minutes * 100 tbr_level_2 = cgm_data[cgm_data['CGM'] < 54].shape[0] * 5 / total_time_minutes * 100 # Adjust TAR level 1 to exclude TAR level 2 counts tar_level_1_adjusted = tar_level_1 - tar_level_2 metrics_list = [num_days_active, percent_data_captured, mean_glucose, gmi, cv, tar_level_2, tar_level_1_adjusted, time_in_range, tbr_level_1, tbr_level_2] metrics_list` |
| Case 5 | `from datetime import datetime # Convert the 'Time' column to datetime format for easier manipulation data['Time'] = pd.to_datetime(data['Time'], format='%m/%d/%y %H:%M') # 1. Number of days the sensor is active num_days = (data['Time'].dt.date.max() - data['Time'].dt.date.min()).days + 1 # Assuming data should be captured every 5 minutes, calculate the total expected measurements # 288 measurements per day (24 hours * 60 minutes / 5 minutes) expected_measurements_per_day = 288 total_expected_measurements = num_days * expected_measurements_per_day # 2. % of sensor data captured percent_data_captured = (len(data) / total_expected_measurements) * 100 # 3. Mean glucose concentration mean_glucose = data['CGM'].mean() # 4. Glucose management indicator (GMI) GMI = 3.31 + 0.02392 * mean_glucose # 5. Glucose variability (coefficient of variation) glucose_cv = (data['CGM'].std() / mean_glucose) * 100 # 6-10. Time in/out of range calculations total_time = len(data) time_above_250 = (data['CGM'] > 250).sum() / total_time * 100 time_above_180 = (data['CGM'] > 180).sum() / total_time * 100 time_in_range = ((data['CGM'] >= 70) & (data['CGM'] <= 180)).sum() / total_time * 100 time_below_70 = (data['CGM'] < 70).sum() / total_time * 100 time_below_54 = (data['CGM'] < 54).sum() / total_time * 100 # Subtract the higher range from the total to get the exclusive percentages for TAR level 1 time_above_180_exclusive = time_above_180 - time_above_250 # List of metrics metrics = [ num_days, percent_data_captured, mean_glucose, GMI, glucose_cv, time_above_250, time_above_180_exclusive, time_in_range, time_below_70, time_below_54 ]` |
|  | metrics |
| Case 6 | `from datetime import datetime # Convert 'Time' column to datetime format for easier manipulation cgm_data['Time'] = pd.to_datetime(cgm_data['Time'], format='%m/%d/%y %H:%M') # Calculate the number of days the sensor was active num_days = (cgm_data['Time'].max() - cgm_data['Time'].min()).days + 1 # Assuming data capture every 5 minutes, calculate % of sensor data captured expected_readings_per_day = 24 * 60 / 5 total_expected_readings = expected_readings_per_day * num_days percent_data_captured = (len(cgm_data) / total_expected_readings) * 100 # Calculate mean glucose concentration mean_glucose = cgm_data['CGM'].mean() # Calculate Glucose Management Indicator (GMI) # GMI formula: 3.31 + (0.02392 * Mean Glucose) gmi = 3.31 + (0.02392 * mean_glucose) # Calculate glucose variability (Coefficient of Variation) cv = cgm_data['CGM'].std() / mean_glucose # Calculate % Time in various glucose ranges total_time = len(cgm_data) time_above_250 = (cgm_data['CGM'] > 250).sum() / total_time * 100 time_above_180 = (cgm_data['CGM'] > 180).sum() / total_time * 100 time_in_range = ((cgm_data['CGM'] >= 70) & (cgm_data['CGM'] <= 180)).sum() / total_time * 100 time_below_70 = (cgm_data['CGM'] < 70).sum() / total_time * 100 time_below_54 = (cgm_data['CGM'] < 54).sum() / total_time * 100 # Adjust time_above_180 to exclude time_above_250 time_above_180_exclusive = time_above_180 - time_above_250 # Create a list with all the metrics metrics = [ num_days, percent_data_captured, mean_glucose, gmi, cv, time_above_250, time_above_180_exclusive, time_in_range, time_below_70, time_below_54 ] Metrics` |
| Case 7 | `from datetime import datetime # Convert 'Time' to datetime format cgm_data['Time'] = pd.to_datetime(cgm_data['Time']) # Calculate the number of days the sensor is active num_days = (cgm_data['Time'].max() - cgm_data['Time'].min()).days + 1 # Assuming the sensor captures data every 5 minutes, calculate total possible readings expected_readings_per_day = 24 * 60 / 5 total_expected_readings = expected_readings_per_day * num_days # Calculate the percentage of sensor data captured percent_captured = (len(cgm_data) / total_expected_readings) * 100 # Calculate mean glucose concentration mean_glucose = cgm_data['CGM'].mean() # Calculate Glucose Management Indicator (GMI) # GMI formula: 3.31 + (0.02392 * mean glucose in mg/dL) gmi = 3.31 + (0.02392 * mean_glucose) # Calculate Glucose Variability (Coefficient of Variation) cv_glucose = cgm_data['CGM'].std() / mean_glucose # Calculate percentages for TAR, TIR, and TBR tar_level_2 = (cgm_data[cgm_data['CGM'] > 250].shape[0] / len(cgm_data)) * 100 tar_level_1 = (cgm_data[cgm_data['CGM'] > 180].shape[0] / len(cgm_data)) * 100 tir = (cgm_data[(cgm_data['CGM'] >= 70) & (cgm_data['CGM'] <= 180)].shape[0] / len(cgm_data)) * 100 tbr_level_1 = (cgm_data[cgm_data['CGM'] < 70].shape[0] / len(cgm_data)) * 100 tbr_level_2 = (cgm_data[cgm_data['CGM'] < 54].shape[0] / len(cgm_data)) * 100 # Adjust TAR Level 1 to exclude TAR Level 2 readings for accurate TAR Level 1 tar_level_1_adjusted = tar_level_1 - tar_level_2 metrics = [ num_days, percent_captured, mean_glucose,` |
|  | `gmi, cv_glucose, tar_level_2, tar_level_1_adjusted, tir, tbr_level_1, tbr_level_2 ] Metrics` |
| Case 8 | `from datetime import datetime # Convert the 'Time' column to datetime format cgm_data['Time'] = pd.to_datetime(cgm_data['Time'], format='%m/%d/%y %H:%M') # Calculate the number of unique days the sensor is active num_days = cgm_data['Time'].dt.date.nunique() # Assuming the sensor should record a reading every 5 minutes, calculate the % of sensor data captured expected_readings_per_day = 24 * 60 / 5 total_expected_readings = expected_readings_per_day * num_days percent_captured = (len(cgm_data) / total_expected_readings) * 100 # Calculate the mean glucose concentration mean_glucose = cgm_data['CGM'].mean() # Calculate the Glucose Management Indicator (GMI) # GMI formula: 3.31 + (0.02392 * mean glucose in mg/dL) gmi = 3.31 + (0.02392 * mean_glucose) # Calculate the glucose variability (coefficient of variation) cv_glucose = (cgm_data['CGM'].std() / mean_glucose) * 100 # Calculate % times in various glucose ranges total_time = len(cgm_data) time_above_250 = (cgm_data['CGM'] > 250).sum() / total_time * 100 time_above_180 = (cgm_data['CGM'] > 180).sum() / total_time * 100 time_in_range = ((cgm_data['CGM'] >= 70) & (cgm_data['CGM'] <= 180)).sum() / total_time * 100 time_below_70 = (cgm_data['CGM'] < 70).sum() / total_time * 100 time_below_54 = (cgm_data['CGM'] < 54).sum() / total_time * 100 # Adjust the % time above range level 1 to exclude time counted in level 2 time_above_180_exclusive = time_above_180 - time_above_250 metrics = [ num_days, # Number of days the sensor is active percent_captured, # % of sensor data captured mean_glucose, # Mean glucose concentration gmi, # Glucose management indicator cv_glucose, # Glucose variability (coefficient of variation) time_above_250, # % Time above TAR in level 2 (>250) time_above_180_exclusive, # % Time above TAR in level 1 (>180) exclusive time_in_range, # % Time in range (70-180mg/dL) time_below_70, # % Time below TBR in level 1 (<70) time_below_54 # % Time below TBR in level 2 (<54) ] metrics` |
| Case 9 | `from datetime import datetime # Convert the "Time" column to datetime format cgm_data['Datetime'] = pd.to_datetime(cgm_data['Time'], format='%m/%d/%y %H:%M') # Calculate the number of days the sensor is active num_days = (cgm_data['Datetime'].max() - cgm_data['Datetime'].min()).days + 1 # Calculate % of sensor data captured # Assuming the sensor records data every 5 minutes, there should be 288 readings per day expected_readings_per_day = 288 total_expected_readings = num_days * expected_readings_per_day percent_captured = (len(cgm_data) / total_expected_readings) * 100 # Calculate mean glucose concentration mean_glucose = cgm_data['CGM'].mean() # Calculate Glucose Management Indicator (GMI) gmi = 3.31 + (0.02392 * mean_glucose) # Calculate glucose variability (coefficient of variation) coefficient_of_variation = cgm_data['CGM'].std() / mean_glucose # Calculate % time above target glucose range (TAR) in level 2 (>250) time_above_250 = len(cgm_data[cgm_data['CGM'] > 250]) / len(cgm_data) * 100` |
|  | `# Calculate % time above target glucose range (TAR) in level 1 (>180) time_above_180 = len(cgm_data[cgm_data['CGM'] > 180]) / len(cgm_data) * 100 # Calculate % time in range (70-180mg/dL) time_in_range = len(cgm_data[(cgm_data['CGM'] >= 70) & (cgm_data['CGM'] <= 180)]) / len(cgm_data) * 100 # Calculate % time below target glucose range (TBR) in level 1 (<70) time_below_70 = len(cgm_data[cgm_data['CGM'] < 70]) / len(cgm_data) * 100 # Calculate % time below target glucose range (TBR) in level 2 (<54) time_below_54 = len(cgm_data[cgm_data['CGM'] < 54]) / len(cgm_data) * 100 # Create a list of the calculated metrics metrics = [ num_days, percent_captured, mean_glucose, gmi, coefficient_of_variation, time_above_250, time_above_180, time_in_range, time_below_70, time_below_54 ] Metrics` |
| Case 10 | `from datetime import timedelta # Parse the 'Time' column to datetime cgm_data['Time'] = pd.to_datetime(cgm_data['Time'], format='%m/%d/%y %H:%M') # Calculate the number of days the sensor is active num_days = (cgm_data['Time'].max() - cgm_data['Time'].min()).days + 1 # Calculate % of sensor data captured (assuming the sensor should capture data every 5 minutes) expected_measurements_per_day = 24 * 60 / 5 total_expected_measurements = expected_measurements_per_day * num_days percent_data_captured = (len(cgm_data) / total_expected_measurements) * 100 # Calculate mean glucose concentration mean_glucose = cgm_data['CGM'].mean() # Glucose Management Indicator (GMI) calculation, using the formula: GMI = 3.31 + 0.02392 * mean glucose gmi = 3.31 + 0.02392 * mean_glucose # Glucose variability as the coefficient of variation (CV = standard deviation / mean * 100) glucose_variability = (cgm_data['CGM'].std() / mean_glucose) * 100 # Calculate % Time in various ranges time_in_tar_level2 = len(cgm_data[cgm_data['CGM'] > 250]) / len(cgm_data) * 100 time_in_tar_level1 = len(cgm_data[(cgm_data['CGM'] > 180) & (cgm_data['CGM'] <= 250)]) / len(cgm_data) * 100 time_in_range = len(cgm_data[(cgm_data['CGM'] >= 70) & (cgm_data['CGM'] <= 180)]) / len(cgm_data) * 100 time_in_tbr_level1 = len(cgm_data[cgm_data['CGM'] < 70]) / len(cgm_data) * 100 time_in_tbr_level2 = len(cgm_data[cgm_data['CGM'] < 54]) / len(cgm_data) * 100 # Time above target range (TAR) >180 includes both level 1 and level 2 time_above_tar = time_in_tar_level1 + time_in_tar_level2 # Compile the results into a list results = [ num_days, # Number of days the sensor is active percent_data_captured, # % of sensor data captured mean_glucose, # Mean glucose concentration gmi, # Glucose management indicator glucose_variability, # Glucose variability (CV) time_in_tar_level2, # % Time above target glucose range (TAR) in level 2 (>250) time_above_tar, # % Time above target glucose range (TAR) in level 1 (>180) including level 2 time_in_range, # % Time in range (70-180mg/dL) time_in_tbr_level1, # % Time below target glucose range (TBR) in level 1 (<70) time_in_tbr_level2 # % Time below target glucose range (TBR) in level 2 (<54) ] Results` |

#### AGP Summarization Task Prompt Output

**Table S9:** Qualitative Prompts Output.

|  |  |
| --- | --- |
| Case | GPT-4 Output (Recorded outputs only) |
| **Case 1** | The CGM data provided covers 14 unique days, which meets the recommended period of analysis. Additionally, the CGM activity time is approximately 88.54%, which is above the recommended threshold of 70%.<br>Output Summary: This data is of good quality because it covers the recommended 14-day analysis period and has a CGM activity time well above the recommended 70% threshold.<br>Hyperglycemia is a periodic concern for this case, with a few days showing higher glucose levels. Notably, January 8th experienced the highest daily percentage of hyperglycemia (12.65%), followed by notable levels on January 13th and 18th. In terms of daily patterns, mornings and afternoons showed higher average glucose readings, with the morning being particularly high. The evening period registered the peak glucose level of 236 mg/dL. However, there were no instances of prolonged severe hyperglycemia, indicating that while there are periods of elevated glucose levels, they do not extend for more than an hour at a time. This information is valuable for identifying specific periods of the day or certain dates where glucose management may require closer attention or intervention.<br>Hypoglycemia was not a prevalent issue for this case. The few instances of hypoglycemia (glucose levels below 70 mg/dL) were observed on January 8th, 14th, and 20th, but these occurrences were minimal and did not constitute a significant part of the day. Notably, there were no instances of severe hypoglycemia (below 54 mg/dL) throughout the analysis period. In terms of daily patterns, a slight increase in hypoglycemic readings was noted during the late night and night, but these did not lead to prolonged events. Overall, the case's glucose levels appear to be well-managed, with no prolonged hypoglycemia episodes, indicating effective control of blood sugar levels to avoid significant low glucose events.<br>The analysis of glycemic variability showed that the case's daily coefficient of variation (CV) consistently remained below the target of 36%, indicating good overall stability in glucose levels. The days with the highest CV, suggesting somewhat greater variability, were January 20th (27.04%), January 14th (27.06%), and January 18th (26.14%), although these values were still within acceptable limits. In terms of the interquartile range (IQR), the highest variability was observed during the evening (53.0 mg/dL) and night (47.0 mg/dL) times. This suggests that glucose levels fluctuate more during these periods compared to other times of the day. Identifying and addressing potential causes for these evening and night fluctuations could further improve the case's glycemic control.<br>Based on the analysis, the primary clinical concern appears to be intermittent hyperglycemia, particularly evident in the morning and afternoon periods, with the highest average glucose levels and occasional spikes above the hyperglycemic threshold. Although the instances of severe hyperglycemia were not prolonged or frequent, identifying and addressing the causes behind these higher glucose levels during specific times of the day could improve overall glycemic control. Hypoglycemia and glycemic variability do not present significant concerns, as the data indicates these aspects are generally well-managed within the recommended guidelines. |
| **Case 2** | The analysis of the CGM data reveals:<br>The data covers 14 unique days, meeting the recommended period of analysis for 14 days.<br>The CGM activity time is approximately 88.81%, which is above the recommended threshold of 70%.<br>Output Summary: This CGM data is of good quality because it covers the recommended analysis period of 14 days and has a high CGM activity time of 88.81%, ensuring a comprehensive and reliable dataset for assessment.<br>Hyperglycemia is a notable concern in this case's CGM data. Interday analysis shows a high variability in glucose levels, with an average of nearly 50% of the day spent in hyperglycemia and peaks reaching as high as 386 mg/dL. Intraday trends reveal specific times of the day, particularly in the evening and night, where hyperglycemia is more pronounced. Critically, there are instances of prolonged severe hyperglycemia, with CGM readings exceeding 250 mg/dL for over an hour. For example, on January 8th, prolonged hyperglycemia was observed from 11:40 to 12:15 and again at 14:30 to 14:45. Similar patterns occurred on January 10th and 19th, indicating the need for targeted interventions during these specific times. These findings suggest the importance of reviewing treatment strategies and lifestyle factors that may influence these patterns.<br>Hypoglycemia is not a predominant issue in this case's CGM data. The analysis shows that the case rarely spends time below the hypoglycemic threshold of 70 mg/dL, both on an interday and intraday basis. Severe hypoglycemia (below 54 mg/dL) was not observed in the data. There was only a single brief instance of prolonged hypoglycemia noted on January 17th between 00:10 and 00:20, but it did not reach the severe hypoglycemia threshold. Overall, the case's glucose levels are well maintained above hypoglycemic levels throughout the majority of the monitoring period, indicating effective management of low glucose levels.<br>The analysis of glycemic variability in the case's data shows overall stability, with all daily coefficients of variation (CV) below the recommended threshold of 36%. The highest CV was observed on January 11th at 30.69%, while the lowest was on January 9th at 19.50%. Regarding the interquartile range (IQR), which reflects glucose fluctuations within specific time windows, the highest variability was noted during the night, with an IQR of approximately 119.75. In contrast, the early morning period exhibited the least variability, with an IQR around 45.00. These findings indicate that while daily glycemic variability is within acceptable limits, there are significant differences in glucose fluctuations across different times of the day, particularly between night and early morning.<br>Based on the analysis and guidelines, the primary clinical concern for this case is hyperglycemia. The data indicates significant periods of both hyperglycemia (glucose levels above 180 mg/dL) and severe hyperglycemia (levels above 250 mg/dL), particularly during certain times of the day and on specific dates. Although hypoglycemia and glycemic variability are not major issues, as evidenced by the minimal occurrences of hypoglycemia and the stable daily coefficients of variation, the consistent and prolonged instances of hyperglycemia warrant clinical attention. It suggests a need for reviewing and potentially adjusting the case's treatment regimen and lifestyle factors to better manage blood glucose levels. |
| **Case 3** | The analysis reveals that:<br>The data covers 14 unique days, meeting the recommended period of analysis.<br>The percentage of CGM activity time is approximately 86.06%, which is above the recommended threshold of 70%.<br>Output Summary: This data is of good quality because it covers the recommended period of analysis (14 days) and has a CGM activity time greater than 70% (86.06%), indicating comprehensive monitoring within the period<br>The analysis of the CGM data indicates that hyperglycemia, including instances of severe hyperglycemia, is a significant concern for this case. Notably, January 10th emerged as a critical day with an average glucose level of 276.21 mg/dL, indicating extensive periods above the hyperglycemia threshold, including 83.97% of readings above 180 mg/dL and 47.39% above 250 mg/dL. Intraday trends revealed that hyperglycemia is particularly prevalent during the afternoon and night, with severe spikes noted on January 10th in the afternoon and January 17th at night. Prolonged hyperglycemia events were identified on several days, including a substantial episode on January 10th spanning from morning to afternoon, and multiple night-time events on January 9th, 17th, and 20th, suggesting challenges in managing glucose levels during these times. These findings highlight the need for targeted interventions to address periods of significant hyperglycemia, especially during identified problematic times of day.<br>The analysis of the CGM data for hypoglycemia shows that it is not a significant issue for this case. A minor instance of hypoglycemia was observed on January 9th, where the glucose level briefly fell to 65 mg/dL during the afternoon, but this did not extend to a prolonged event or reach severe hypoglycemia levels. There were no instances of glucose levels falling below 54 mg/dL, and no prolonged periods of hypoglycemia exceeding 60 minutes were identified. This suggests that the case's current management strategies for preventing hypoglycemia are generally effective, with only a singular, brief deviation noted. Monitoring should continue, but there appears to be no immediate cause for concern |
|  | regarding hypoglycemia based on this data analysis.<br>The glycemic variability analysis revealed that several days exhibited high variation in glucose levels, notably on January 9th, 10th, 17th, and 20th, where the coefficient of variation exceeded the guideline of 36%. This indicates significant fluctuations in glucose levels on these days, suggesting that factors affecting glycemic control may need closer examination and management. Furthermore, the interquartile range (IQR) analysis identified the evening as the time of day with the highest glucose fluctuations, followed by the afternoon and night. The least variability was observed during the early morning hours. These findings highlight the need for targeted strategies to manage glucose levels more effectively, particularly during times of day identified with higher variability, to improve overall glycemic control for the case.<br>Based on the analysis, the primary clinical concern is the significant hyperglycemia, particularly the instances of severe hyperglycemia and high glycemic variability. Days with notable hyperglycemia, such as January 10th and 17th, and times of day like the evening and afternoon, were identified as periods of high glucose levels and variability. The occurrence of prolonged hyperglycemia events further underscores the urgency of addressing these glycemic control issues. While hypoglycemia does not appear to be a significant problem for this case, the high levels of variability and instances of severe hyperglycemia suggest that the current diabetes management strategy may need to be reevaluated and adjusted to better control blood glucose levels and minimize risks associated with hyperglycemia. |
| Case 4 | The CGM data analysis reveals the following:<br>The dataset covers 14 unique days, meeting the recommended period of analysis.<br>The CGM activity time is approximately 90.3%, which is above the recommended threshold of 70%.<br>Output Summary: This data is of good quality because it encompasses the recommended 14 days of analysis and has a CGM activity time well above the recommended threshold, ensuring a comprehensive and reliable dataset for further analysis.<br>The analysis of the CGM data indicates that hyperglycemia is a significant issue for the case, with several days showing a high percentage of time spent above the hyperglycemia threshold. Notably, on January 9, 11, 15, 16, 17, 18, and 19, the case spent a large part of the day in a hyperglycemic state, with January 15 showing an exceptionally high 98.6% of the time above the threshold. Severe hyperglycemia (>250 mg/dL) was also a concern, particularly on January 12 and 18, where more than half of the day was spent at these dangerous levels. Prolonged hyperglycemia events were frequent, with significant episodes occurring across various days, including multiple instances on January 9, 12, 15, 18, and 19 in different time windows such as the afternoon, morning, and late night. These findings suggest the need for a detailed review of the case's treatment plan and lifestyle choices during these identified periods to address and mitigate hyperglycemia risks effectively.<br>The analysis of the CGM data for hypoglycemia reveals that low blood sugar levels are not a significant concern for the case during the observed period. Only on January 10th was there a minor incidence of hypoglycemia (below 70 mg/dL), with a negligible portion of the day affected and no evidence of severe hypoglycemia (below 54 mg/dL) on any day. Furthermore, there were no instances of prolonged hypoglycemia, indicating that the case did not experience extended periods of dangerously low glucose levels. This lack of significant hypoglycemia suggests that while managing high blood sugar levels is crucial for the case, maintaining a balance to avoid overly aggressive lowering that could result in hypoglycemia is also being effectively achieved.<br>The glycemic variability analysis revealed that most days exhibited a coefficient of variation (CV) below the recommended threshold of 36%, indicating generally stable glucose levels. However, specific days, such as January 10, 12, and 18, demonstrated higher CVs, with January 18 approaching the threshold, suggesting notable fluctuations in glucose levels. The interquartile range (IQR) analysis highlighted January 12 and 18 as days with significantly larger fluctuations, indicative of pronounced glycemic variability. Furthermore, the analysis of IQR across different times of the day identified "Late night" and "Afternoon" as periods with the highest variability, pointing towards more substantial fluctuations during these times. This information could be crucial for tailoring case management strategies, particularly focusing on dietary and activity adjustments during times of heightened variability to enhance glycemic control.<br>The primary clinical concern identified from the analysis is the significant issue of hyperglycemia, with several days showing a high percentage of time spent above the hyperglycemia threshold and instances of severe hyperglycemia. This is compounded by the observation of pronounced glycemic variability on certain days and at specific times, suggesting that while hypoglycemia is not a significant issue, the case's glucose management strategy may need adjustment. The frequent and prolonged periods of high glucose levels, especially the severe hyperglycemia events, pose a considerable risk for diabetic complications. Therefore, addressing these hyperglycemic episodes and managing glycemic variability should be a priority in the case's diabetes care plan. |
| Case 5 | The CGM data covers 14 unique days, meeting the recommended period of analysis. Additionally, the CGM activity time is approximately 97.67%, which is well above the recommended threshold of 70%.<br>Output Summary: The data is of good quality because it covers the recommended 14-day analysis period and has a high CGM activity time of approximately 97.67%, exceeding the recommended 70%.<br>The analysis of your CGM data indicates that hyperglycemia is not a significant concern. Interday trends show that on average, only about 0.63% of your time is spent in a hyperglycemic state (glucose > 180 mg/dL), with the highest daily percentage being around 5.21%. Intraday trends reveal that your average glucose levels vary throughout the day but do not frequently exceed the hyperglycemia threshold, with the highest recorded level being 208 mg/dL. Importantly, there were no instances of prolonged severe hyperglycemia (glucose levels > 250 mg/dL lasting over an hour) in the data. These findings suggest good overall glucose management, although it might be beneficial to review the days with slightly higher levels to understand any specific triggers or activities associated with these variations.<br>The analysis of your CGM data reveals that hypoglycemia is not a significant concern. Interday trends show minimal daily occurrences of hypoglycemia, with very low percentages of time spent below both the mild (below 70 mg/dL) and severe (below 54 mg/dL) hypoglycemic thresholds. Intraday trends do not indicate any consistent pattern or specific times of day when hypoglycemia is more likely to occur. Importantly, there were no instances of prolonged hypoglycemia, either mild or severe, on any of the days in the analyzed dataset. This suggests effective management of your glucose levels, with minimal risk of hypoglycemic episodes.<br>The analysis of glycemic variability in your CGM data reveals excellent glucose control. No days were identified with a coefficient of variation exceeding the recommended 36%, indicating a consistent and stable management of glucose levels on a daily basis. Furthermore, the interquartile ranges for different times of the day – Morning, Afternoon, Evening, and Night – were consistently low, suggesting minimal variability in glucose levels throughout the day. This stability in glycemic control is a positive indicator of effective diabetes management strategies, and it minimizes the risk of complications associated with large glucose fluctuations.<br>Based on the analysis of the CGM data and adherence to clinical guidelines, the primary clinical concern for the case appears to be minimal. The case's glucose levels are well-managed, with minimal instances of hyperglycemia or hypoglycemia. Glycemic variability is also within the recommended range, indicating stable glucose control. Therefore, the case seems to be effectively managing their diabetes, and there are no significant clinical concerns identified from this data analysis. However, continued monitoring and routine check-ups are advisable to maintain this level of control. |
| Case 6 | Duration of Data Collection: The dataset contains CGM readings for exactly 14 unique days, meeting the recommended period of analysis for assessing diabetes management.<br>CGM Activity Time: The dataset has a CGM activity time of approximately 92.4%, which is significantly above the recommended threshold of 70%. |
|  | Output Summary<br>The data is of good quality because it covers the recommended 14-day analysis period and has a high CGM activity time of over 92%, ensuring a comprehensive overview of the case's glucose levels.<br>Hyperglycemia is a significant concern for this case, with both frequent and severe instances noted. Interday analysis revealed particularly high glucose levels on specific days, such as January 21 and 13, where over 70% of readings were above the hyperglycemia threshold. Intraday trends showed the highest glucose peaks during late night and night, indicating a pattern of elevated glucose levels during these periods. Prolonged hyperglycemic events, where glucose levels were above 250 mg/dL for an hour or more, were observed on several days, including the afternoon and night of January 9, and the early morning and morning of January 10. These findings highlight the need for a focused review of the case's diabetes management, especially during the identified times and days.<br>Hypoglycemia is generally not a major concern for this case, with only a few isolated instances. The most notable occurrence was on January 20, 2023, when 6.6% of the day's readings were below the hypoglycemia threshold of 70 mg/dL, particularly during late night. This specific instance indicates a need for cautious nighttime glucose management. Intraday analysis revealed that the lowest glucose readings typically occurred during the late-night hours, though severe hypoglycemia (below 54 mg/dL) was not a prominent issue. There was one prolonged hypoglycemic event on January 20, highlighting the importance of monitoring and possibly adjusting treatment strategies during late-night periods. Overall, while hypoglycemia is not widespread, vigilance during late-night hours is advisable.<br>"The analysis of glycemic variability within the dataset revealed significant fluctuations in glucose levels on specific days and during certain times of the day. Notably, January 18 and January 9 exhibited high coefficients of variation, 44.54% and 42.41% respectively, surpassing the recommended threshold of 36%. This indicates considerable daily variability in glucose levels on these days, suggesting the need for closer monitoring or adjustments in diabetes management. Regarding the interquartile range, the most significant variability in glucose readings occurred during late night (IQR of 112) and evening (IQR of 102), indicating these periods as the times with the highest fluctuations. These findings underscore the importance of focusing on stabilizing glucose levels, particularly during late nights and evenings, to manage glycemic variability effectively.<br>Based on the analysis and guidelines, the primary clinical concern for this case is the significant hyperglycemia, including severe hyperglycemia, with frequent high glucose levels and substantial daily variability. Particular attention should be given to managing elevated glucose levels during late night and evening hours, as well as on days like January 18 and 9, which showed high glycemic variability. Although hypoglycemia is not a widespread issue, occasional monitoring, especially during late-night hours, is advisable. The focus should be on stabilizing blood glucose levels to avoid extreme highs and manage variability effectively. |
| Case 7 | Based on the analysis:<br>The CGM data covers 14 unique days, which meets the recommended period of analysis for 14 days.<br>The CGM was active approximately 95.49% of the time, well above the recommended activity time of 70%.<br>Output Summary: The data is of good quality as it covers the recommended 14 days and has a high CGM activity time of about 95.49%.<br>Hyperglycemia is a significant issue for this case, with several days showing elevated glucose levels. Notably, January 9th and 18th were particularly concerning, with extremely high percentages of the day spent in severe hyperglycemia. Intraday trends indicate that morning glucose levels are consistently high, followed by elevated levels in the afternoon and evening. Prolonged severe hyperglycemia events were frequent; for instance, on January 8th, these events spanned across the afternoon, evening, morning, and night. Such patterns suggest the need for careful review and potential adjustments in diabetes management, especially considering the severity and frequency of these hyperglycemic episodes.<br>Hypoglycemia is generally not a significant concern for this case, as the data shows very few instances of low glucose levels. The only notable exception occurred on January 16th during late night, where both hypoglycemia (below 70 mg/dL) and severe hypoglycemia (below 54 mg/dL) were observed for a short duration. This isolated event of prolonged hypoglycemia, lasting over 60 minutes for hypoglycemia and over 15 minutes for severe hypoglycemia, warrants attention to understand possible triggers or specific conditions on that night. Otherwise, the case's glucose levels were mostly stable and above the hypoglycemic thresholds.<br>Output Summary: The glycemic variability analysis reveals that most days exhibit stable glucose levels with a Coefficient of Variation (CV) below the recommended 36%. However, increased variability was noted on January 13th, 15th, 16th, and particularly on January 21st, where the CV exceeded the threshold, indicating more fluctuating glucose levels. The Daily Interquartile Range (IQR) analysis showed that mornings and nights are periods with the highest glucose variability, with mornings having an average IQR around 51.7 and nights around 59.1. These findings suggest that while the case's overall glycemic control is relatively stable, there are specific times, particularly mornings and nights, where glucose levels demonstrate more significant fluctuations.<br>Based on the analysis, the primary clinical concern for this case is the frequent and severe hyperglycemia. Several days, notably January 9th and 18th, showed extremely high levels of glucose and a significant portion of time spent in severe hyperglycemia. Additionally, the case experienced substantial glucose variability on certain days and during specific times (especially mornings and nights), which could complicate diabetes management. While hypoglycemia does not appear to be a major issue, the high and variable hyperglycemic episodes are a significant concern and should be addressed with the case's healthcare provider to optimize their diabetes treatment plan. |
| Case 8 | The data covers 14 unique days and has a CGM activity time of approximately 91.5%.<br>Output Summary:<br>This data is of good quality because it meets the recommended analysis period of 14 days and has a CGM activity time greater than 70%, specifically around 91.5%.<br>Hyperglycemia is a noticeable concern for this case, with evidence of glucose levels exceeding the hyperglycemia threshold of 180 mg/dL on several days, though no instances of severe hyperglycemia (over 250 mg/dL) were recorded on a daily aggregate level. The data did not show any prolonged severe hyperglycemia events, indicating that while glucose levels frequently rise above desired levels, they do not remain at critically high levels for extended periods. Intraday analysis revealed varying glucose levels throughout the day, suggesting that certain times may be more prone to hyperglycemic episodes; however, without severe spikes reaching the threshold for prolonged concern. This information can be useful for identifying patterns in daily activities or dietary habits that may influence glucose levels. It is important for the case to be aware of these trends to manage and mitigate hyperglycemic episodes more effectively.<br>The analysis of the CGM data for hypoglycemia indicates that this case does not experience significant issues with low blood sugar levels. No days were identified with a concerning number of readings below the hypoglycemia or severe hypoglycemia thresholds, suggesting that episodes of low glucose are not dominating the case's glucose management. Furthermore, there were no instances of prolonged hypoglycemia, where glucose levels stayed dangerously low for extended periods. This lack of prolonged hypoglycemic events is a positive sign, indicating effective management of glucose levels to avoid the risks associated with low blood sugar. It suggests that the case's current diabetes management plan is effectively preventing severe hypoglycemia, a crucial goal in the treatment of diabetes.<br>The analysis of glycemic variability within the CGM data revealed no days with a coefficient of variation exceeding 36%, indicating that the case maintains relatively stable |
|  | glucose levels on a daily basis without significant day-to-day fluctuations. This stability suggests effective management of glycemic variability, a crucial factor in minimizing the risk of diabetes-related complications. The interquartile range (IQR), which measures the spread of the middle 50% of glucose readings, varied across different times of the day, with a mean IQR of approximately 26.75 mg/dL. Although there are some fluctuations in glucose levels throughout the day, these did not show extreme variability, indicating controlled glucose levels during different times. This analysis underscores the importance of ongoing monitoring and management to maintain this level of glycemic control and stability.<br>Based on the analysis, the primary clinical concern appears to be hyperglycemia, with glucose levels frequently exceeding the recommended upper limit of 180 mg/dL, though not reaching severe hyperglycemia levels above 250 mg/dL for prolonged periods. While there are no significant concerns regarding hypoglycemia or excessive glycemic variability, managing and reducing episodes of hyperglycemia remains a crucial aspect of the case's diabetes management. Identifying and addressing the causes of these hyperglycemic episodes, whether related to diet, physical activity, or insulin therapy adjustments, should be a focus to minimize the risk of diabetes-related complications. |
| Case 9 | The dataset contains data for 14 unique days, and the CGM was active for approximately 99.16% of the time.<br>Given this information, we can conclude that the data is of good quality because:<br>It covers the recommended period of analysis of 14 days.<br>The CGM activity time is well above the recommended threshold of 70%.<br>Output Summary: This CGM data is of good quality because it spans over the recommended 14 unique days and has a high activity time of approximately 99.16%, indicating comprehensive monitoring.<br>The analysis of the CGM data reveals significant concerns regarding hyperglycemia for this case, particularly on certain days and during specific times. Notably, January 12th and 13th stood out with exceptionally high average glucose levels and significant portions of the day spent above the hyperglycemic threshold, indicating severe hyperglycemia. Additionally, there were multiple instances of prolonged severe hyperglycemia, notably during the late night and early morning of January 12th, and during the afternoon and evening of January 13th. These patterns suggest that the case's glucose levels tend to rise significantly during the late night to early morning and in the afternoon to evening. Identifying specific behaviors or circumstances on these days and times, such as diet, exercise, or medication management, could be crucial in developing strategies to mitigate these hyperglycemic events.<br>The hypoglycemia analysis indicates that while hypoglycemic events were not widespread across all days, January 11th emerged as a significant concern due to notable levels of hypoglycemia, including both below 70 mg/dL for a substantial part of the evening and severe hypoglycemia below 54 mg/dL. Additionally, prolonged hypoglycemia events were observed on January 11th and 12th, particularly in the evening and night, suggesting a pattern where the risk of hypoglycemia increases during these times. These findings highlight the importance of closely monitoring glucose levels and possibly adjusting treatment or behaviors around these identified times to mitigate the risk of future hypoglycemic episodes.<br>The glycemic variability analysis highlighted significant fluctuations in glucose control, particularly on January 11th, 12th, and 13th, where the coefficient of variation exceeded the target of less than 36%, indicating high glycemic variability. These days, especially with CVs of 43.86%, 55.17%, and 53.41% respectively, suggest inconsistent glucose management and potential risk areas for the case. Additionally, the evening time window exhibited the highest interquartile range (IQR) of glucose readings, pointing to a broader variability in glucose levels during this period. This suggests that the case's glucose levels are more unstable in the evenings, possibly due to dinner meals, activity levels, or the timing of medication. Addressing these specific times and patterns could be key to achieving more stable glucose control.<br>The primary clinical concern based on the analysis is the significant glycemic variability, particularly the instances of severe hyperglycemia and the days with high coefficient of variation, indicating inconsistent glucose control. The days with prolonged severe hyperglycemia (glucose levels above 250 mg/dL for extended periods) and the evenings with high interquartile ranges of glucose readings highlight the need for closer monitoring and possibly adjustments in the management plan. These findings suggest that the case may benefit from tailored interventions to address both hyperglycemia and glycemic variability, potentially involving changes in insulin therapy, dietary adjustments, and lifestyle modifications to stabilize glucose levels and reduce the risk of complications. |
| Case 10 | The CGM data covers 14 unique days with a span of 14 days, and the CGM activity time is approximately 97.37%. This means the dataset meets both recommended criteria for analysis:<br>It has 14 unique days of data, aligning with the recommended period of analysis.<br>The CGM was active about 97.37% of the time, which is well above the recommended threshold of 70%.<br>Output Summary: The data is of good quality because it covers the recommended period of 14 days and has a high CGM activity time of approximately 97.37%, ensuring a comprehensive analysis<br>The analysis of the CGM data indicates that hyperglycemia is a concern for this case, particularly with several instances of severe hyperglycemia. Notably, the evenings of January 11th and the afternoons of January 12th and January 18th exhibited significant hyperglycemia, with glucose levels peaking at 285 mg/dL, 271 mg/dL, and 332 mg/dL, respectively. These findings suggest a pattern of postprandial hyperglycemia that might be linked to meal composition or timing. Prolonged periods of severe hyperglycemia, lasting for an hour or more, were identified on January 8th (early hours), January 11th (late afternoon and early evening), January 12th (early afternoon), and January 18th (midday and early afternoon), indicating critical times when the case's glucose management was inadequate. These specific instances provide actionable insights for the case and their healthcare provider to explore potential triggers and adjust treatment plans accordingly.<br>The analysis of the CGM data reveals that hypoglycemia is a notable concern for this case, particularly with occurrences of prolonged and severe hypoglycemic events. Notably, the nights transitioning from January 11th to 12th and the early morning of January 16th experienced significant periods of hypoglycemia below 70 mg/dL for more than an hour, and instances where glucose levels dropped below 54 mg/dL for more than 15 minutes. These findings suggest a pattern of nocturnal hypoglycemia, which could be related to insulin dosing, meal planning, or physical activity levels. Addressing these trends is crucial for preventing hypoglycemia and ensuring the case's safety, indicating a need for potential adjustments in the case's diabetes management strategy, especially in terms of nighttime insulin dosing or snack recommendations.<br>The analysis of glycemic variability in the CGM dataset reveals an overall stable pattern, with an average daily coefficient of variation (CV) at 24.10%, indicating good glycemic control for most days. However, there was a day where the CV spiked to 43.08%, highlighting a day with significantly higher glycemic variability, which may warrant closer examination to understand potential causes such as dietary choices, activity levels, or insulin dosing. The interquartile range (IQR) analysis shows higher variability during the night, early morning, and afternoon times, with the night time window exhibiting the highest IQR at 50.00 mg/dL. These findings suggest that these periods are particularly prone to fluctuations in blood glucose levels, and targeted strategies may be needed to address this variability and enhance the case's glycemic stability.<br>The primary clinical concern based on the analysis is the presence of both significant hyperglycemia and hypoglycemia events, indicating challenges in maintaining stable glycemic control. Specifically, the occurrence of severe hyperglycemia, with glucose readings frequently exceeding 250 mg/dL, and instances of prolonged hypoglycemia below 70 mg/dL, suggest risks for both acute and long-term complications. Additionally, the variability in glucose levels, especially during specific times of the day, points to the need for adjustments in the case's diabetes management plan. Addressing these fluctuations through tailored interventions could help mitigate risks and improve overall glycemic stability. |

#### AGP Summarization Task Prompt Code Written

**Table S10:**
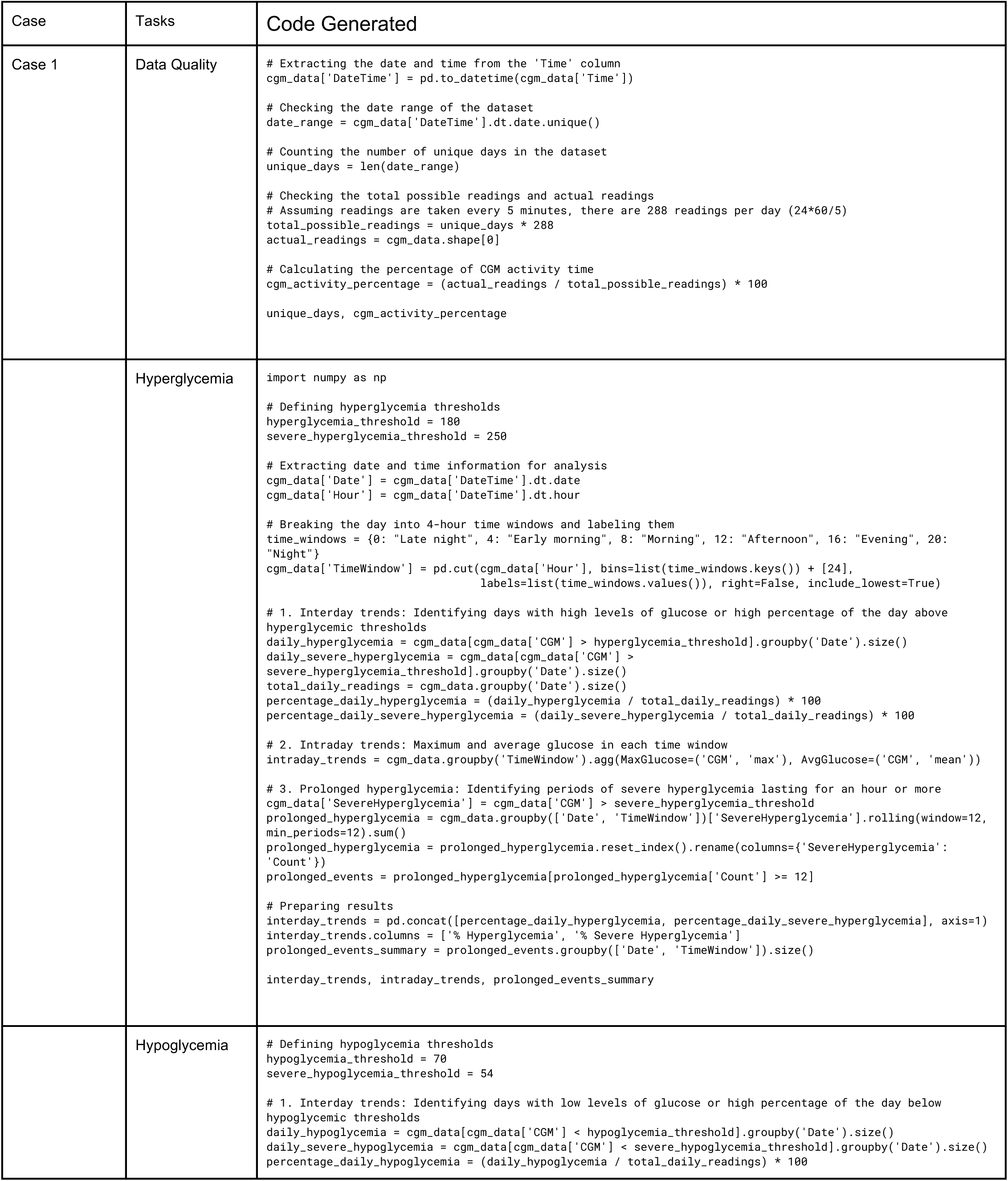
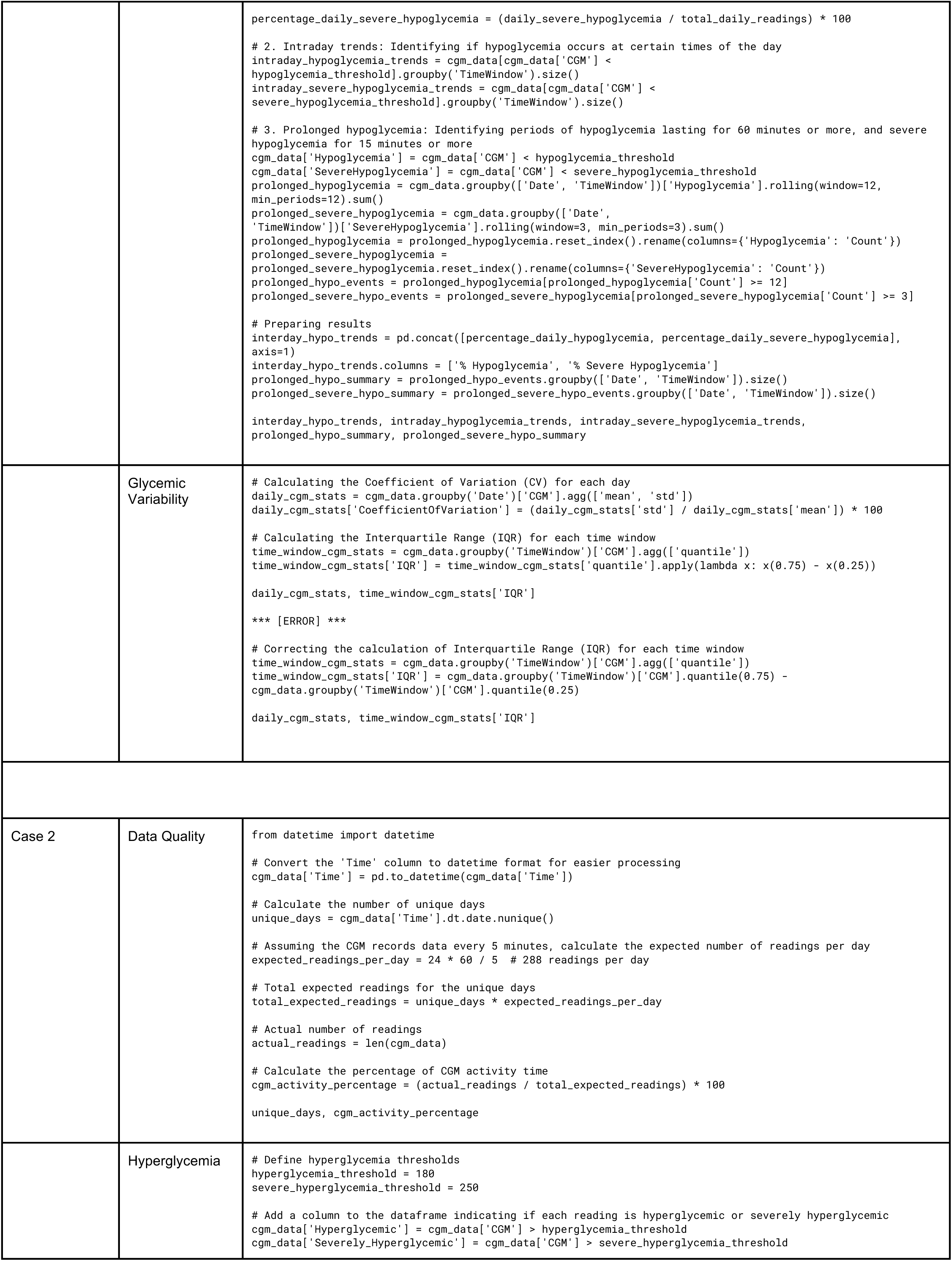
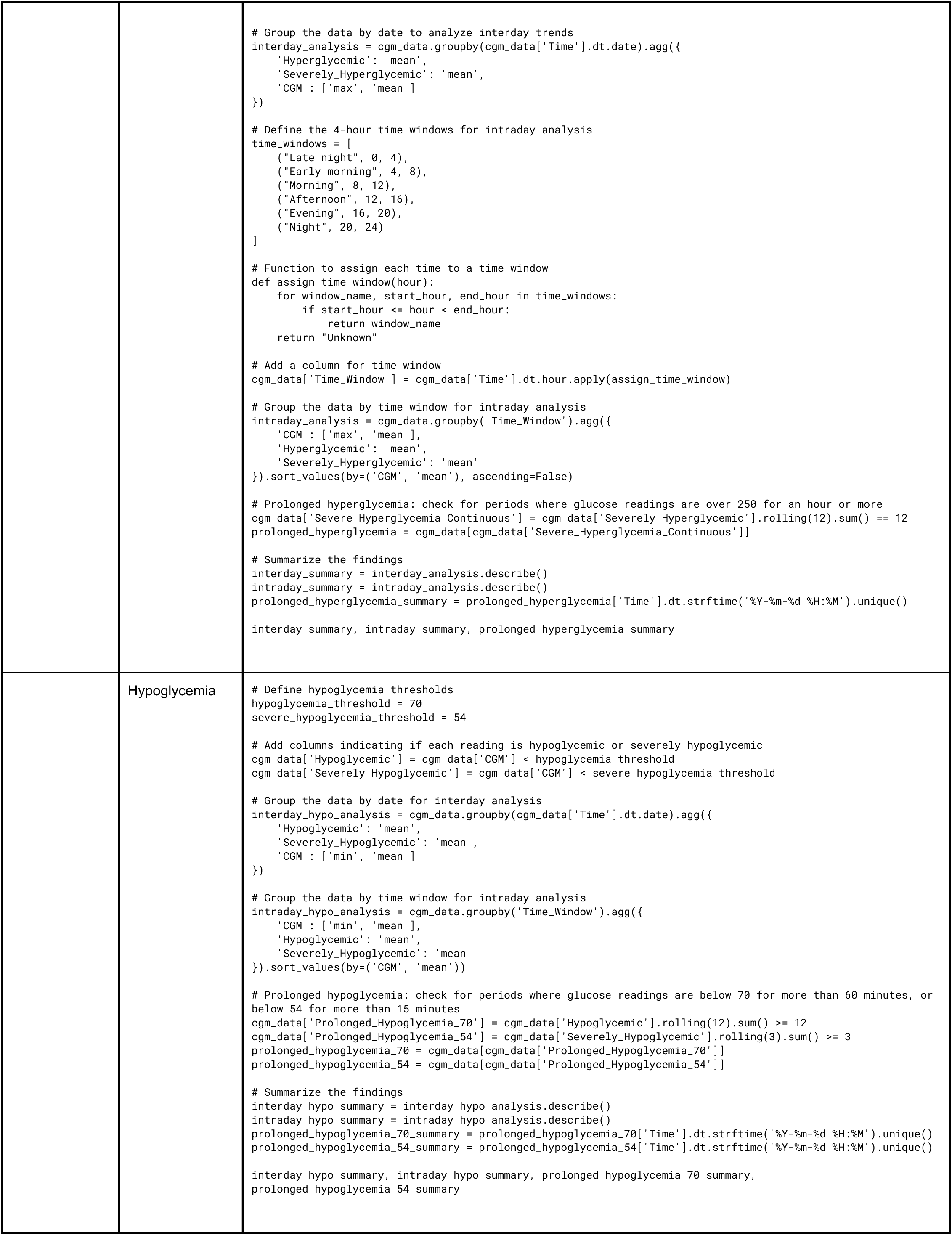
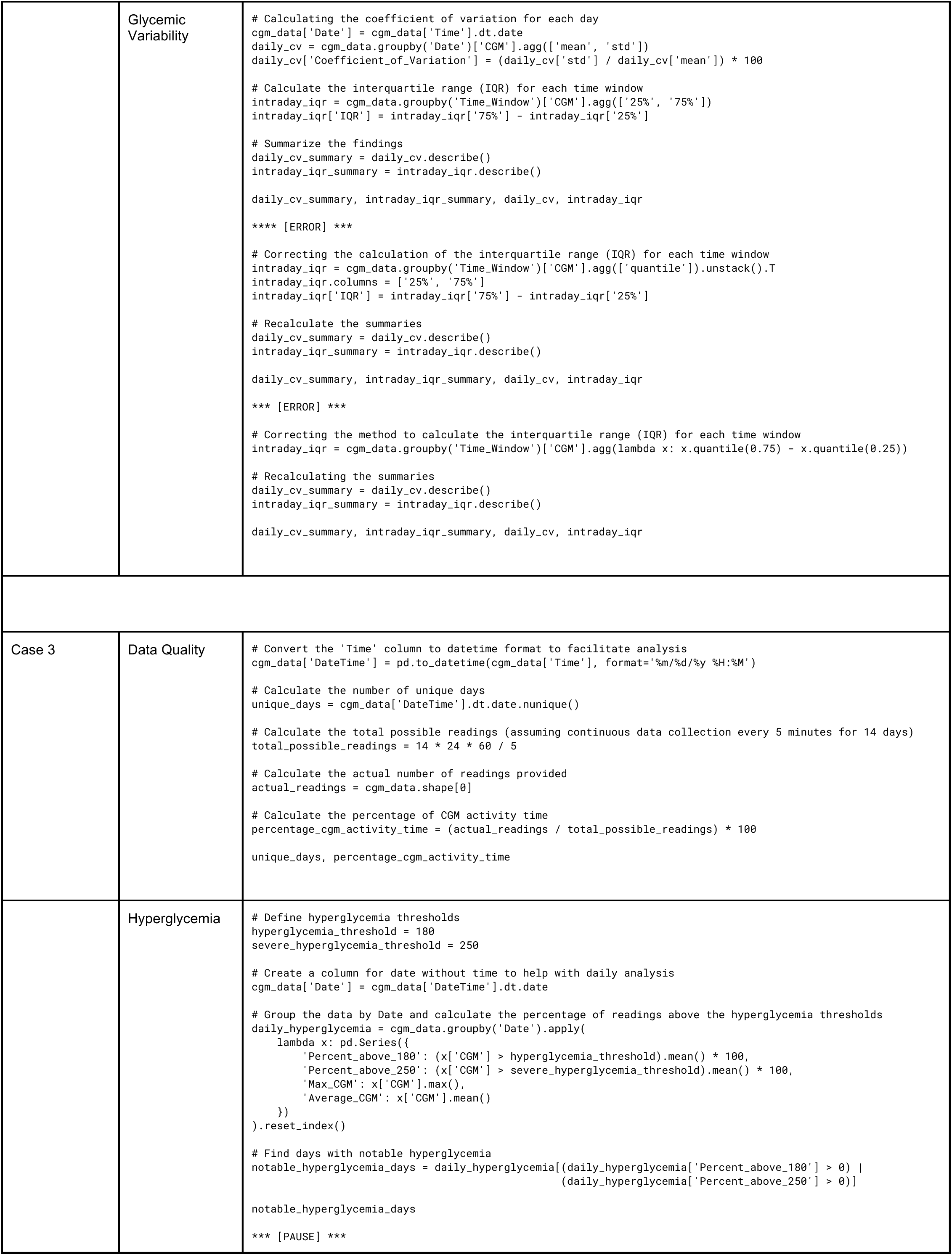
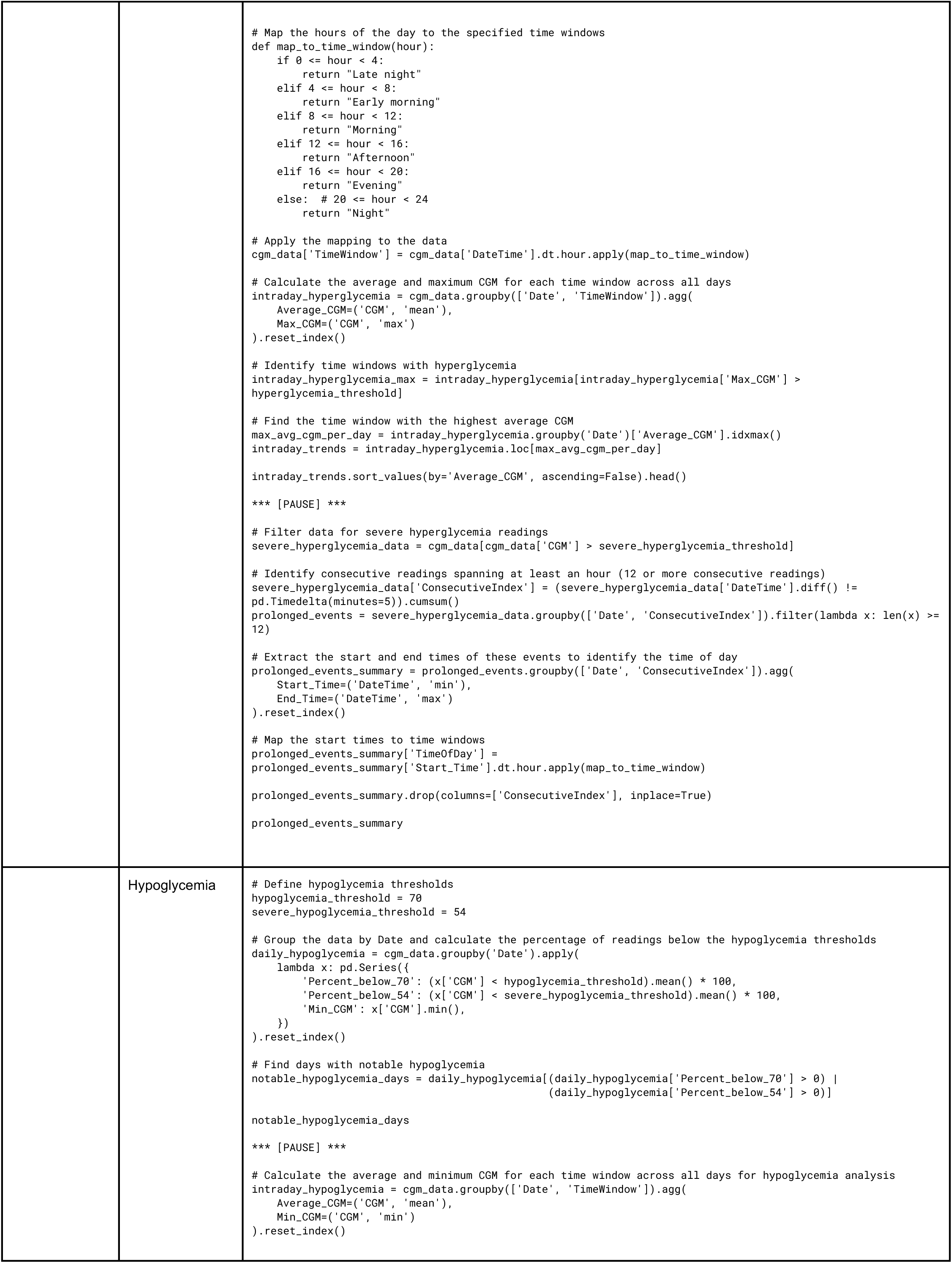
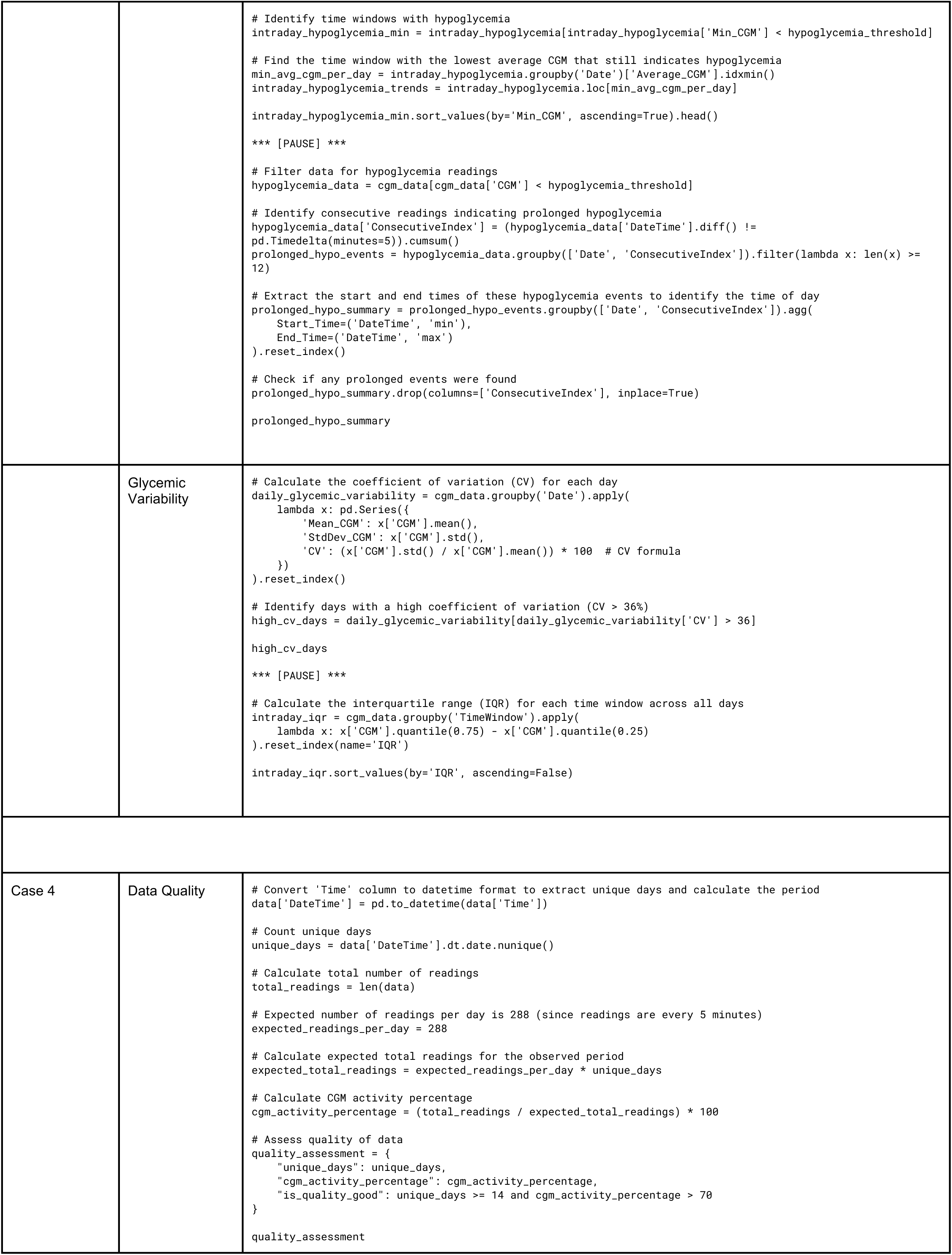
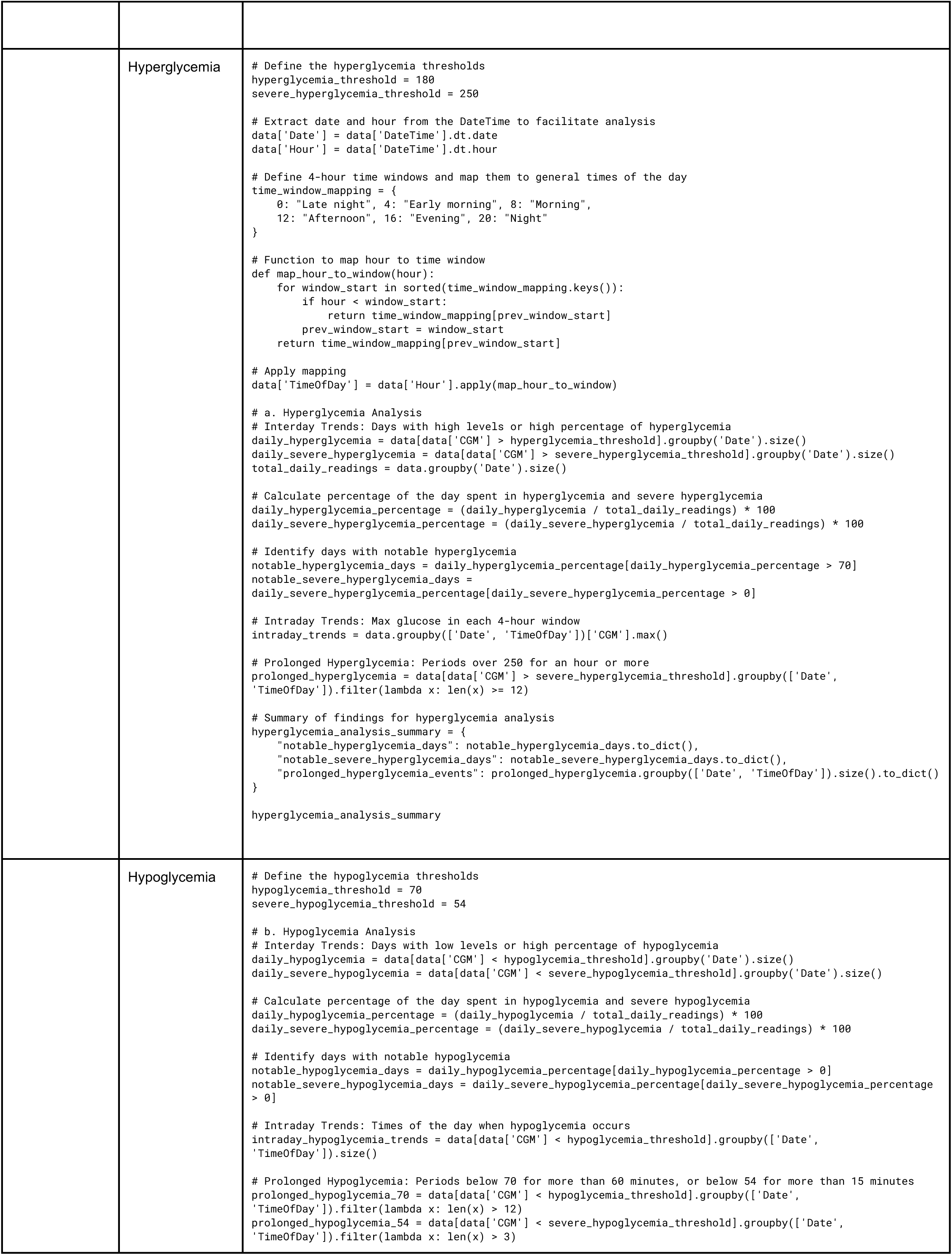
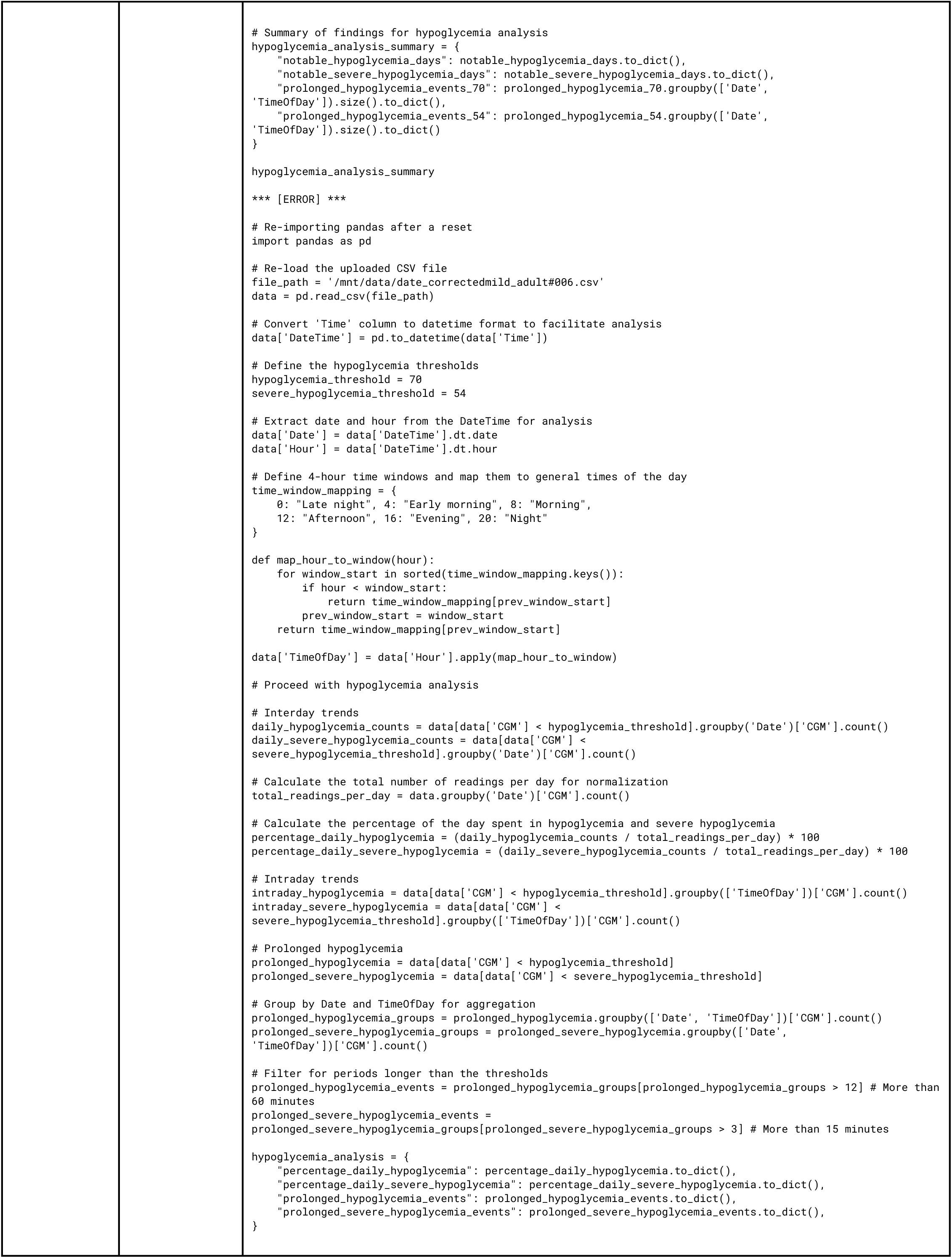
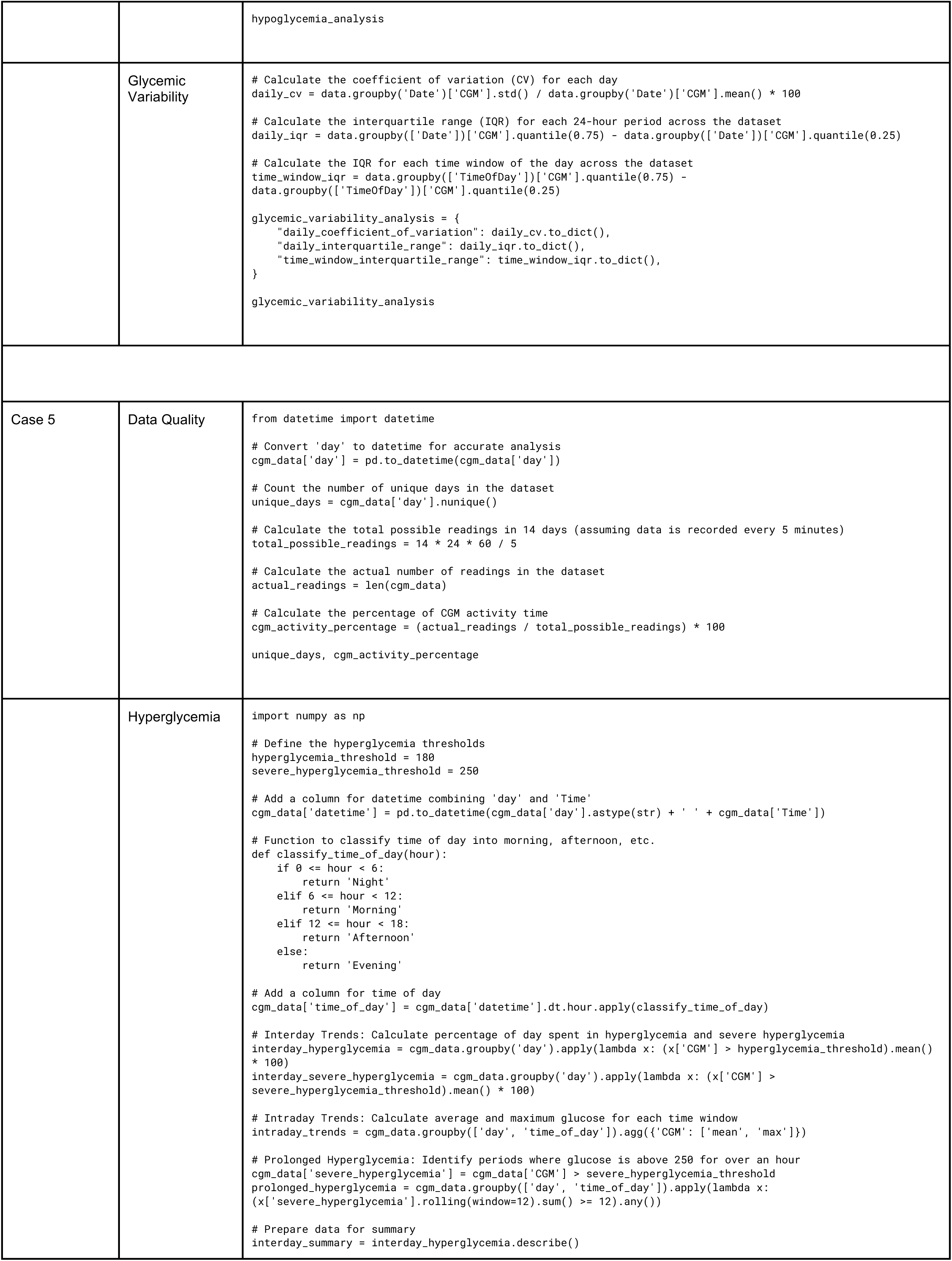
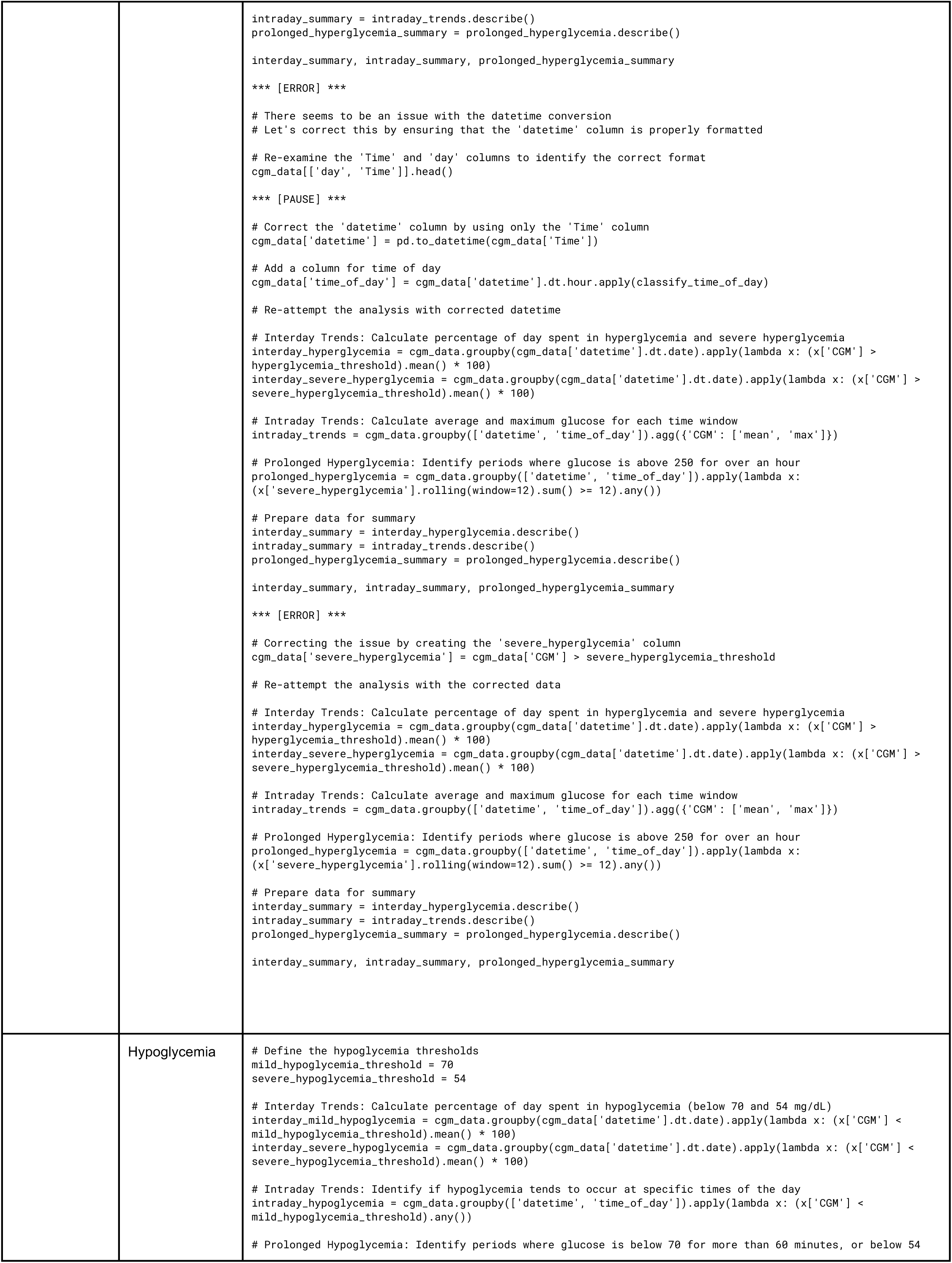
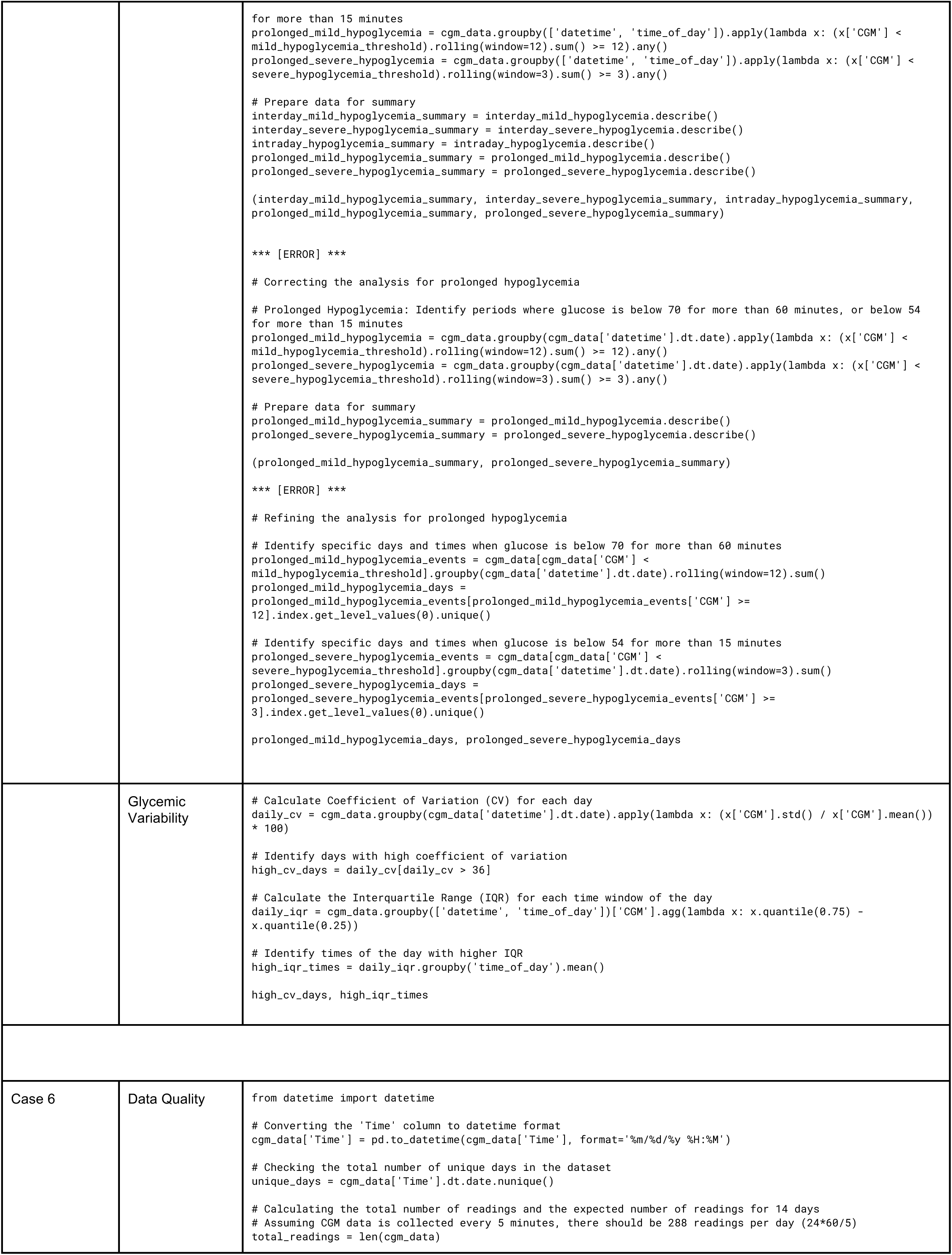
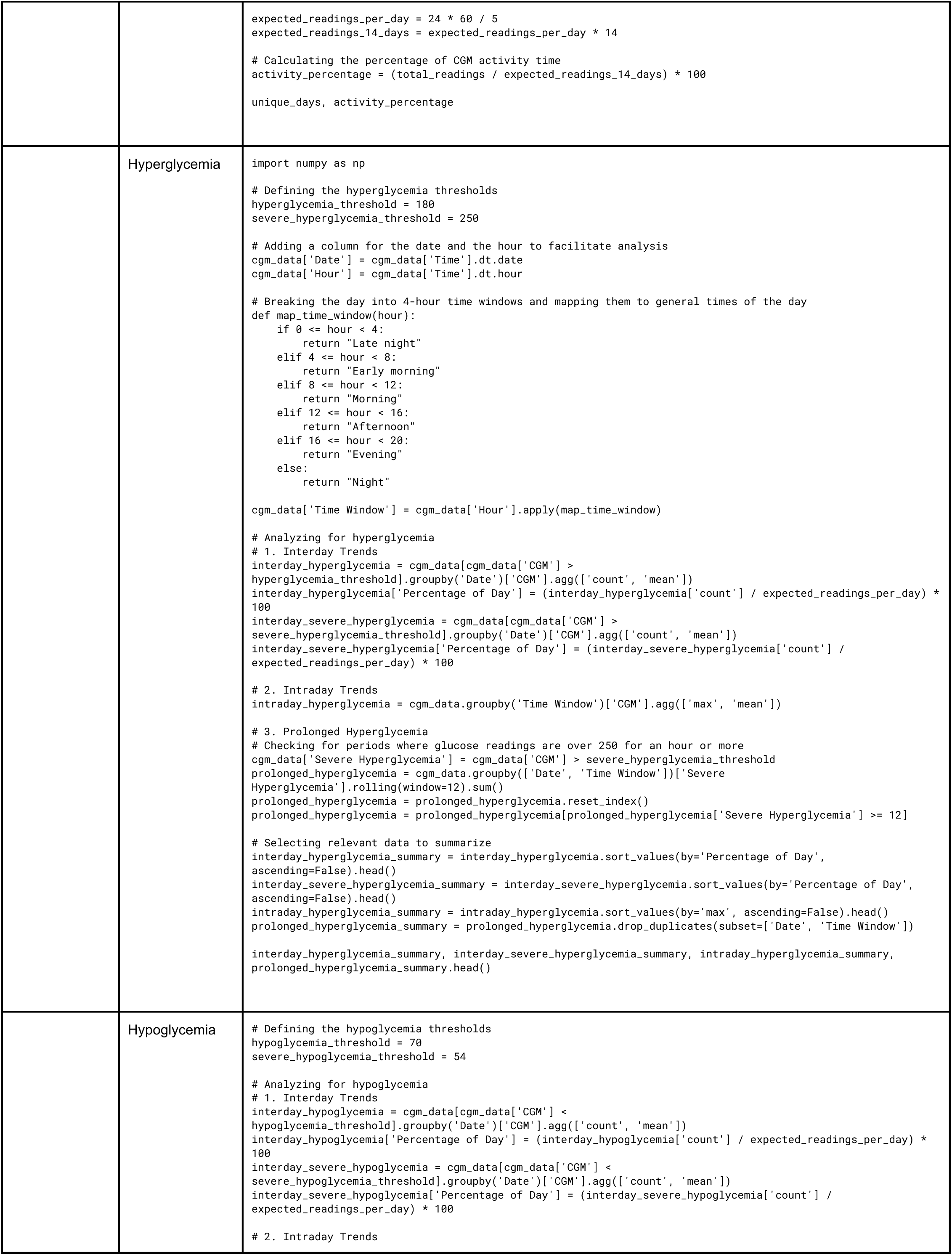
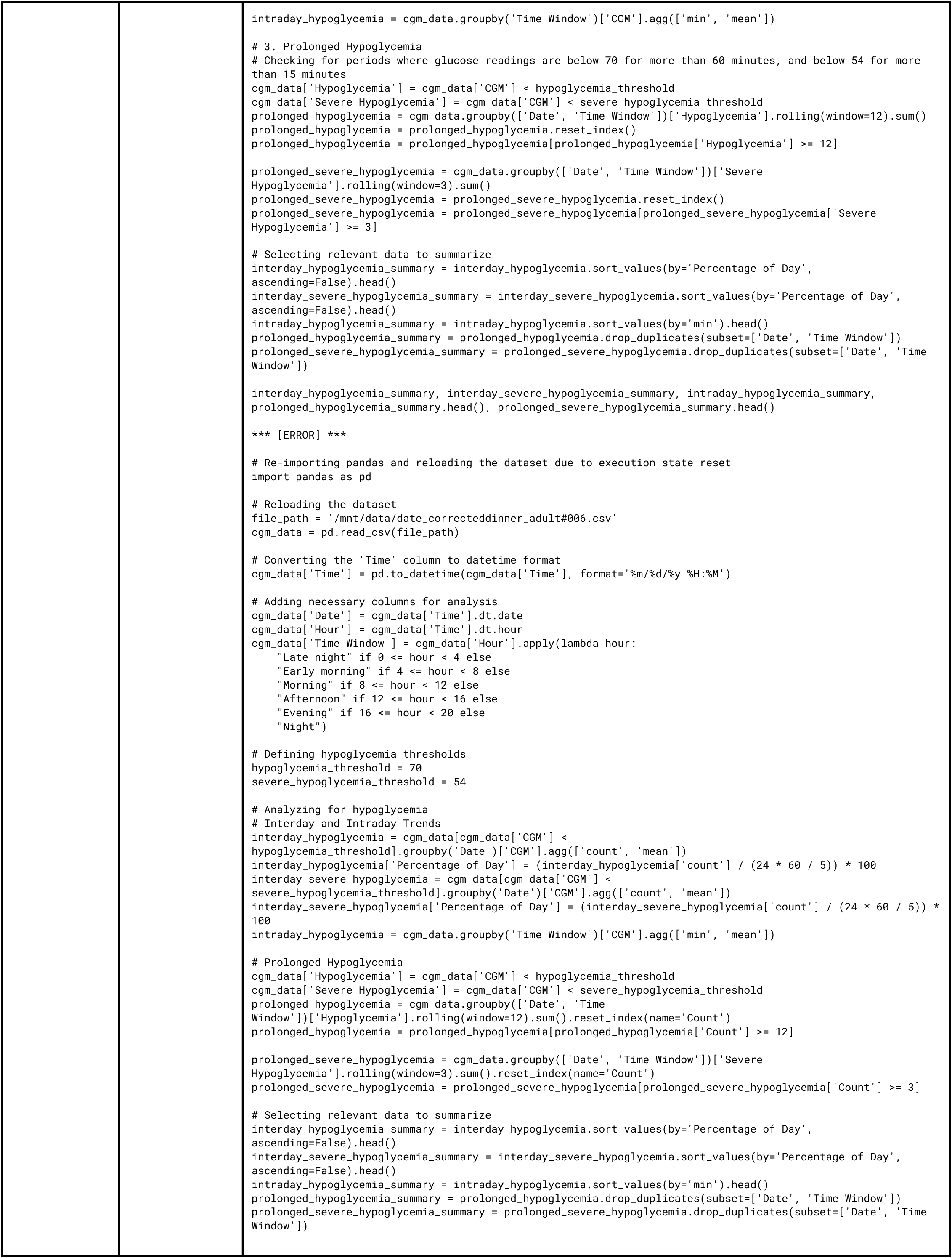
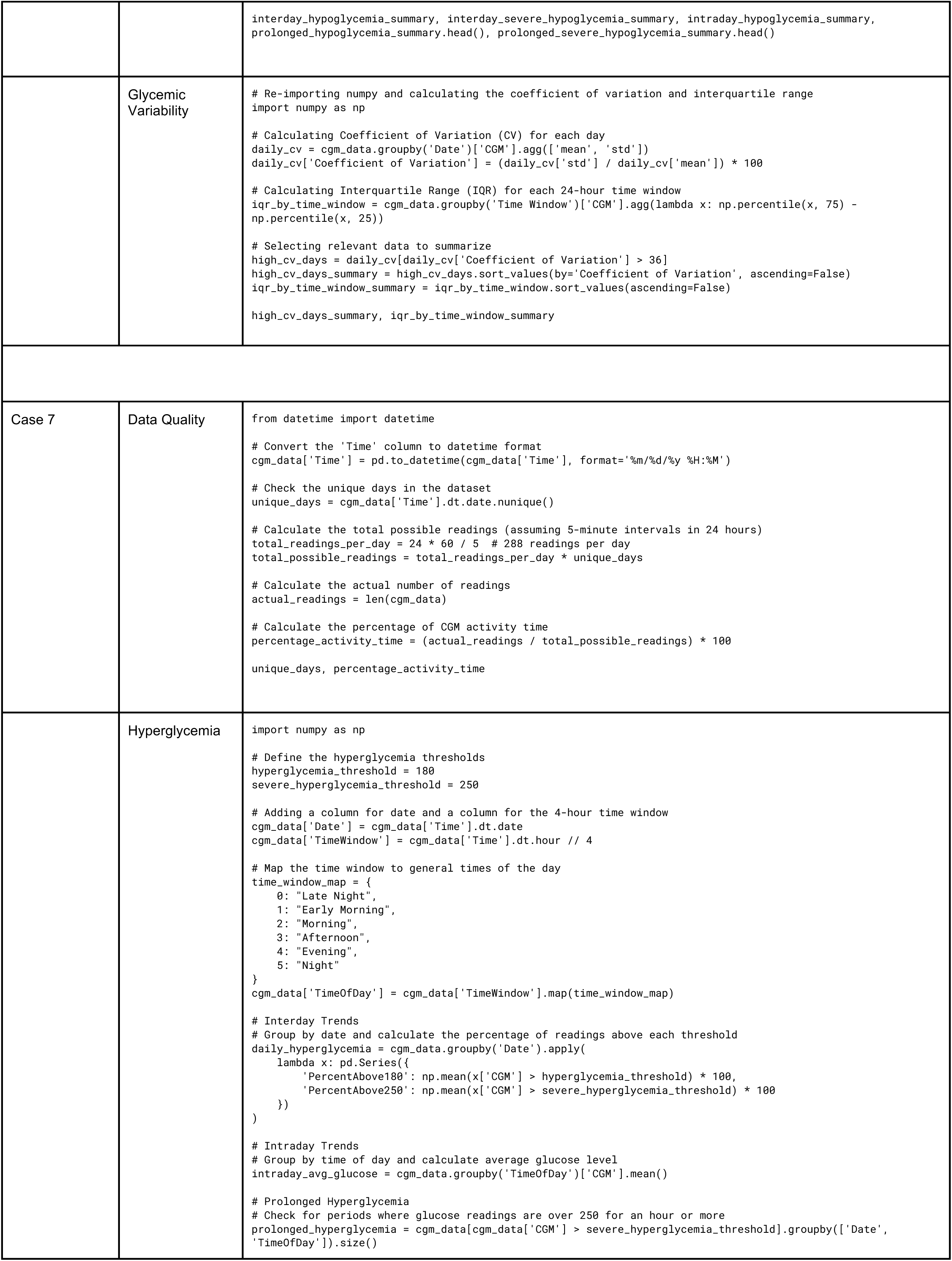
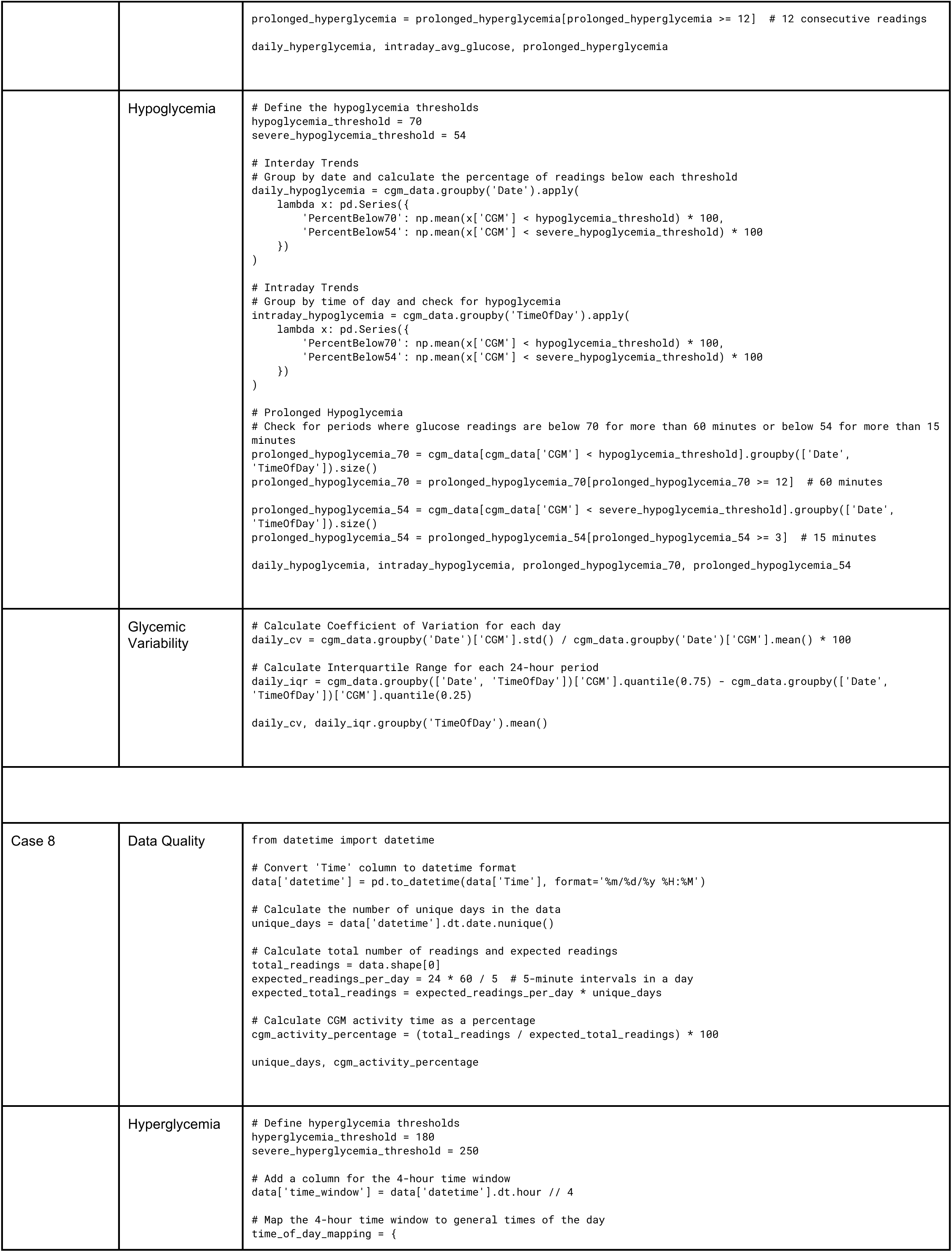
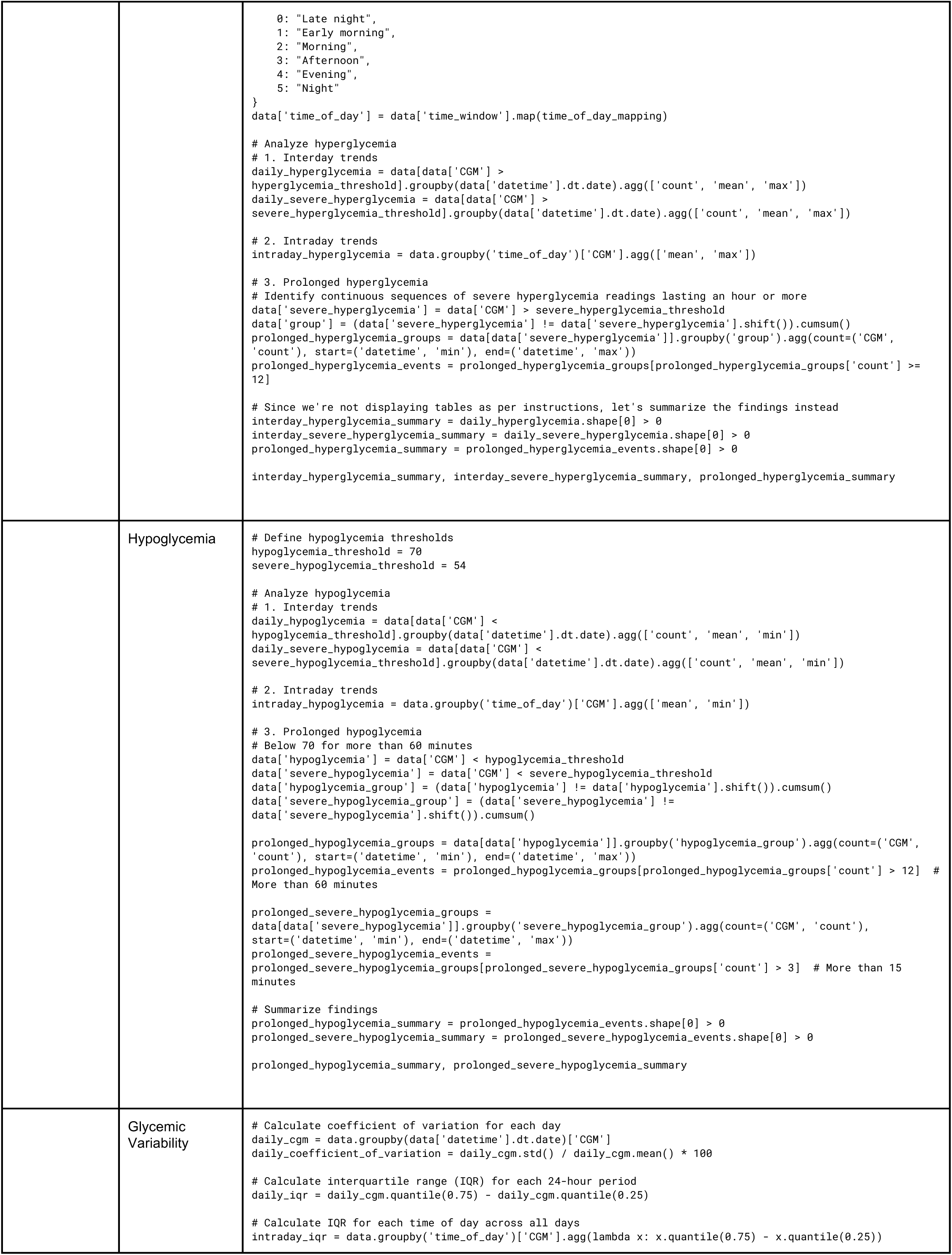
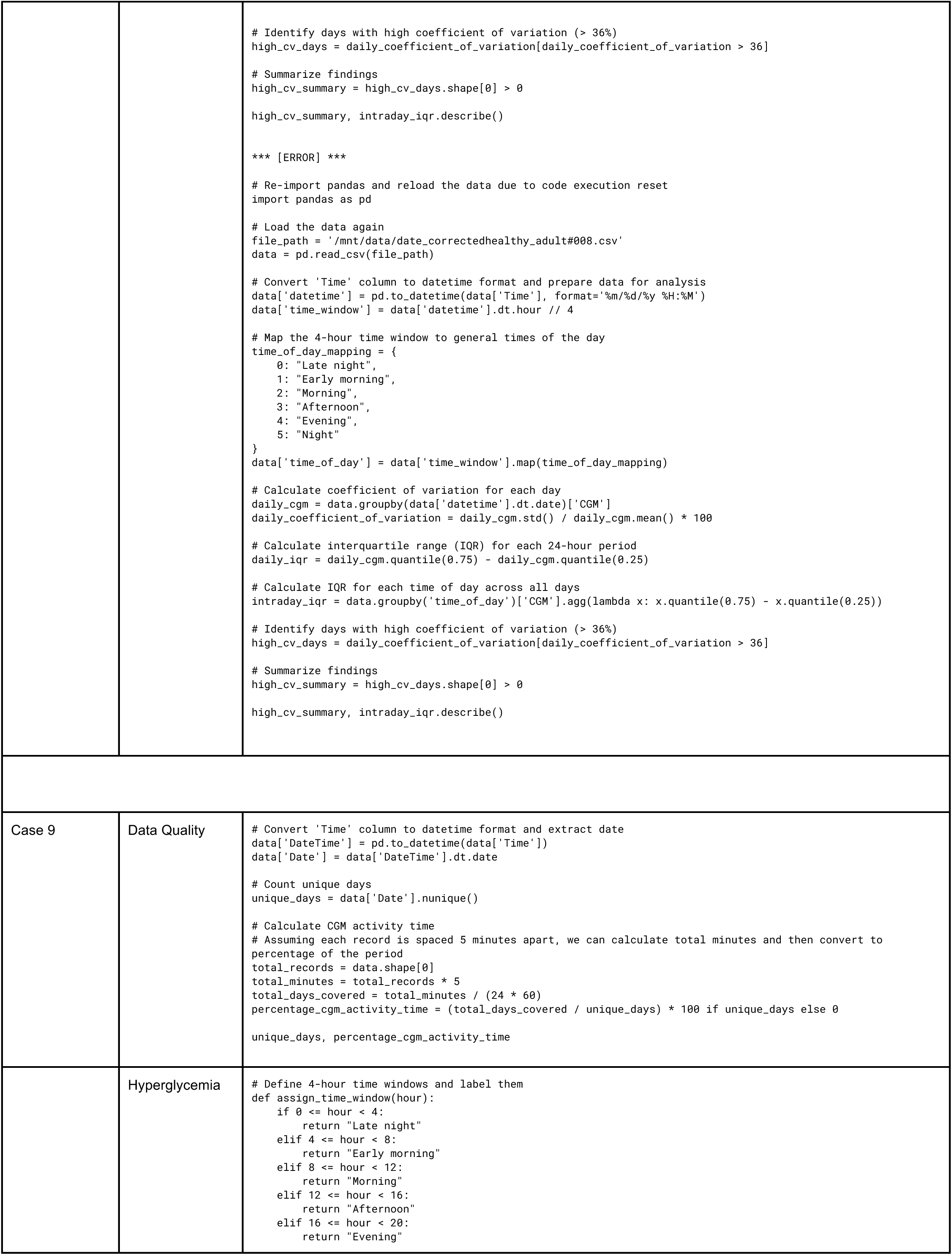
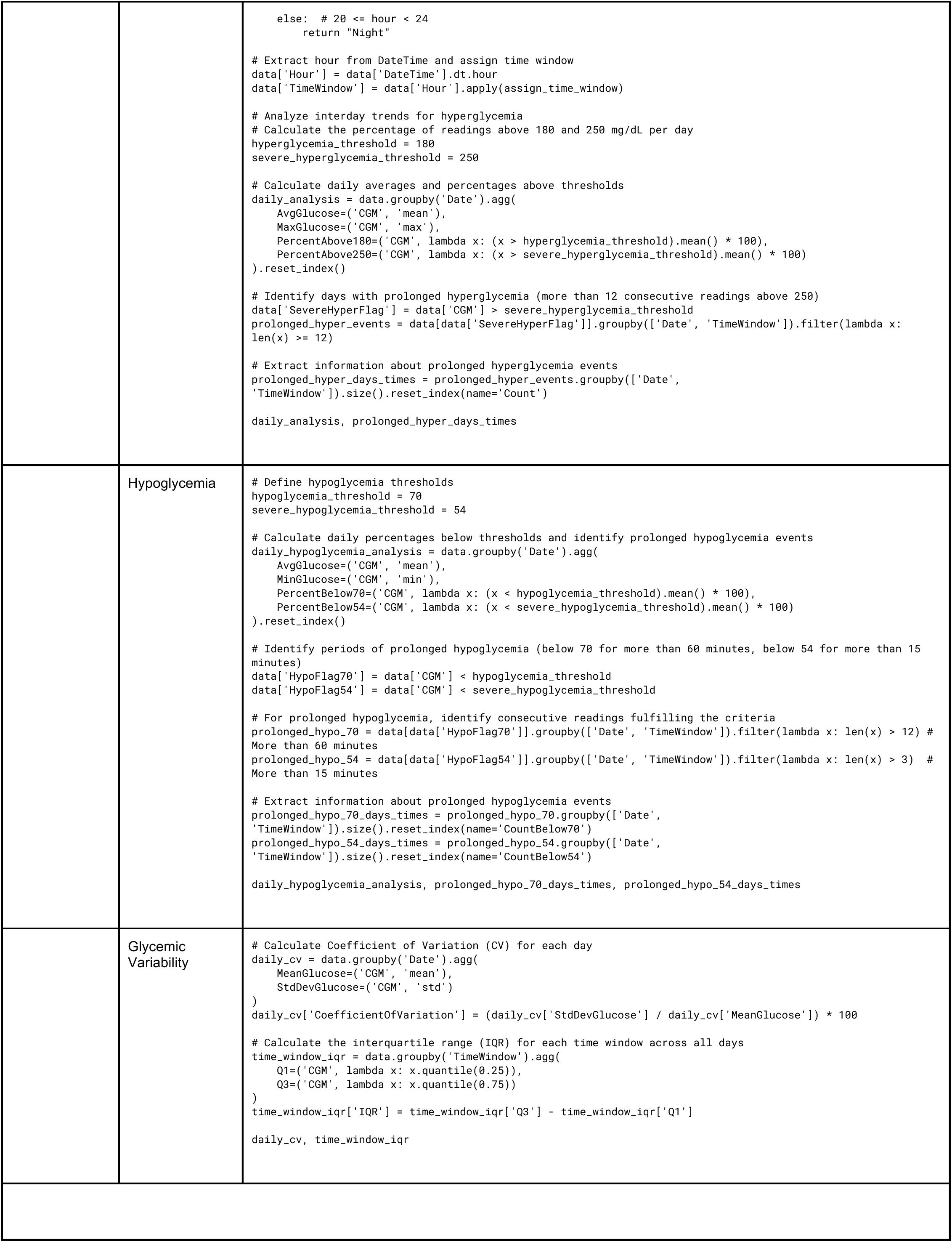
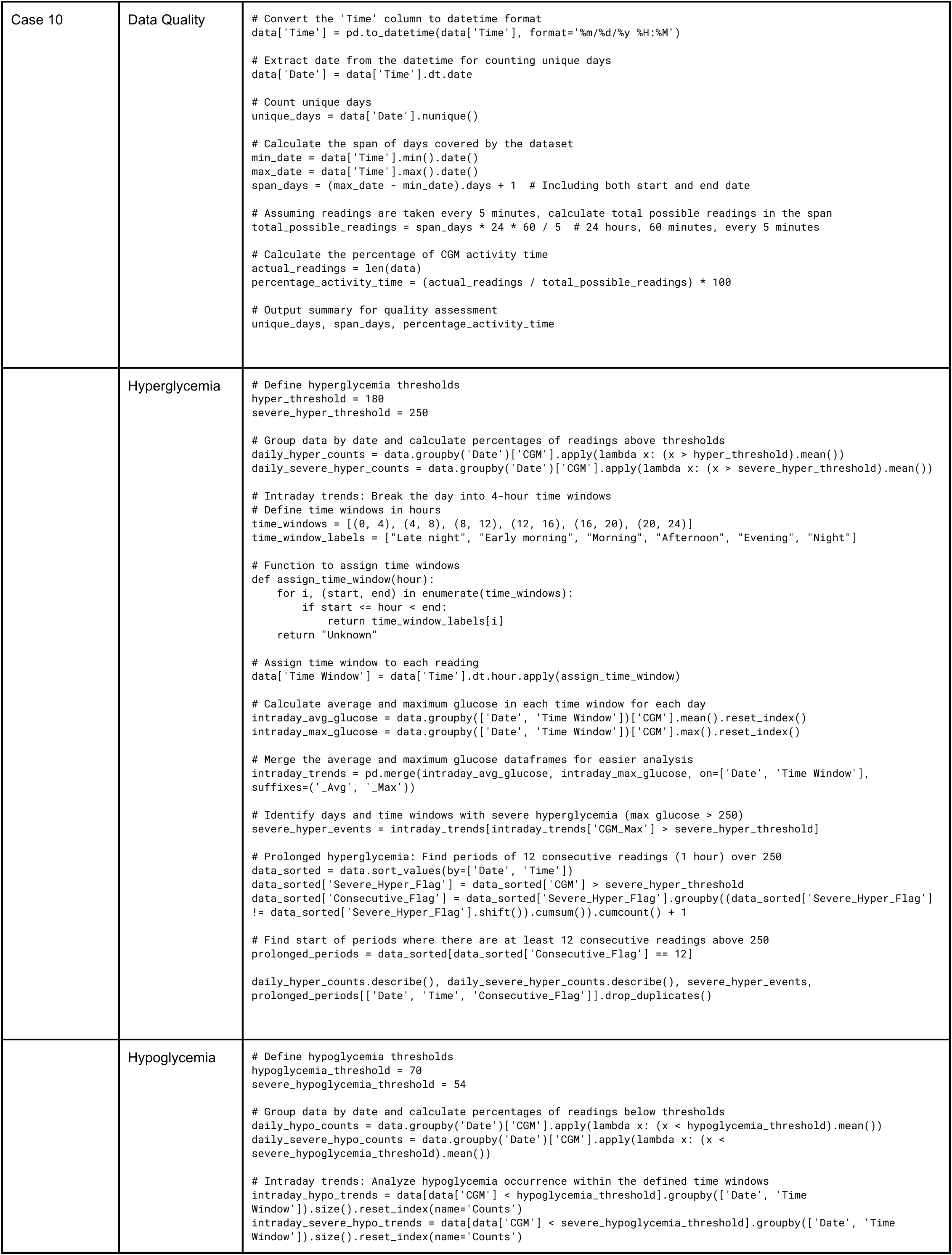
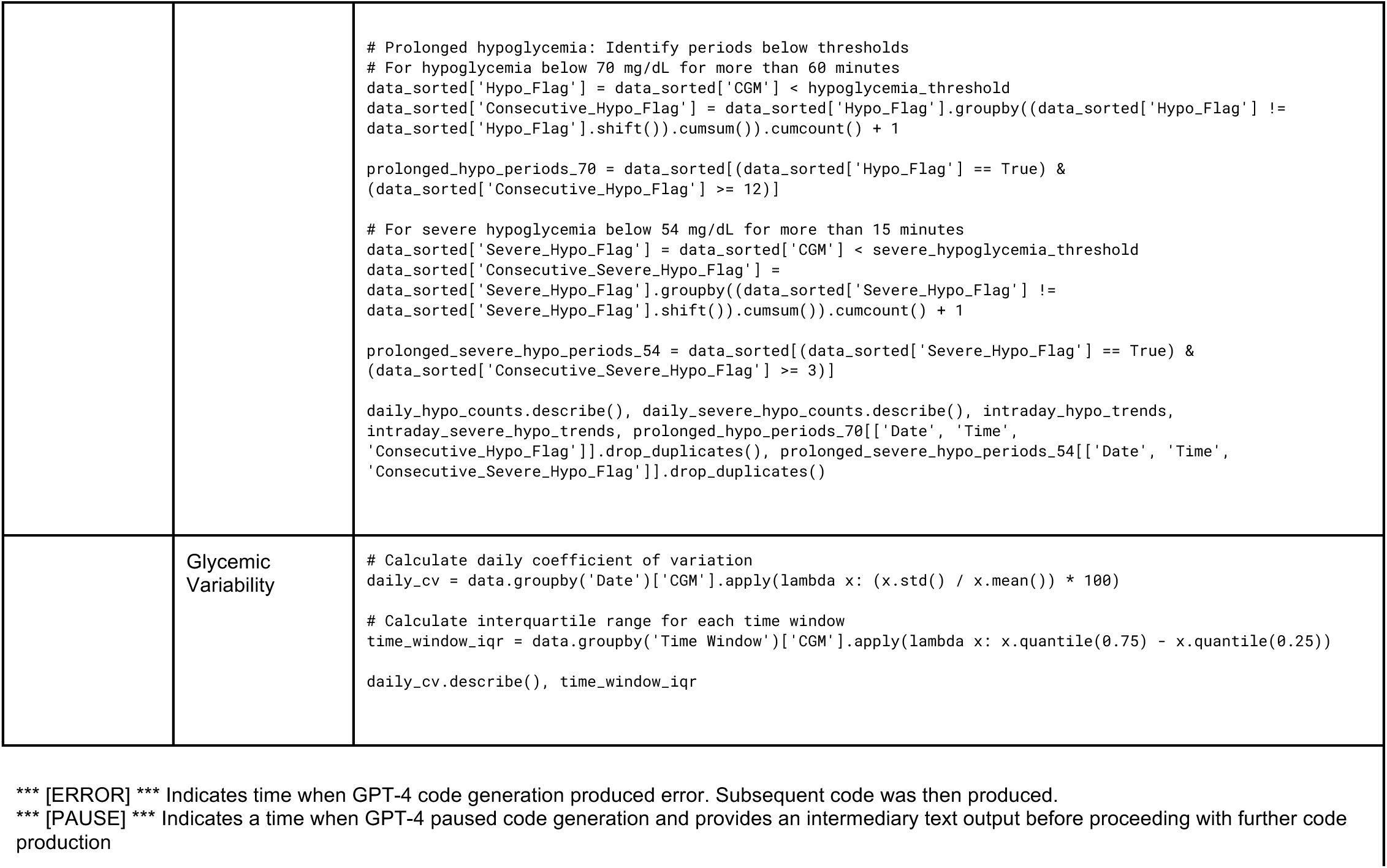
Qualitative Prompts Code written by GPT-4.

